# Influenza vaccine effectiveness against influenza-associated hospitalizations and emergency department or urgent care encounters among children and adults — United States, 2024–25 season

**DOI:** 10.64898/2026.04.22.26350853

**Authors:** Jennifer DeCuir, Emily L. Reeves, Zachary A. Weber, Duck-Hye Yang, Stephanie A. Irving, Sara Y. Tartof, Nicola P. Klein, Shaun J. Grannis, Toan C. Ong, Sarah W. Ball, Malini B. DeSilva, Kristin Dascomb, Allison L. Naleway, Padma Koppolu, S. Bianca Salas, Lina S. Sy, Bruno Lewin, Richard Contreras, Ousseny Zerbo, John R. Hansen, Lawrence Block, Karen B. Jacobson, Brian E. Dixon, Colin Rogerson, Thomas Duszynski, William F. Fadel, Michelle A. Barron, David Mayer, Catia Chavez, Adam Yates, Lindsey Kirshner, Charlene E. McEvoy, Omobosola O. Akinsete, Inih J. Essien, Tamara Sheffield, Daniel Bride, Julie Arndorfer, Josh Van Otterloo, Karthik Natarajan, Caitlin S. Ray, Amanda B. Payne, Katherine Adams, Brendan Flannery, Shikha Garg

## Abstract

**Background:** The 2024–25 influenza season was the most severe in the United States (US) since 2017–18, with co-circulation of both influenza A virus subtypes (H1N1 and H3N2). Influenza vaccine effectiveness (VE) has varied by season, setting, and patient characteristics.

**Methods:** Using electronic healthcare encounter data from eight US states, we evaluated influenza vaccine effectiveness (VE) against influenza-associated hospitalizations and emergency department or urgent care (ED/UC) encounters from October 2024–April 2025 among children aged 6 months–17 years and adults aged ≥18 years. Using a test-negative, case-control design, we compared the odds of influenza vaccination between acute respiratory illness (ARI) encounters with a positive (cases) versus negative (controls) test for influenza by molecular assay, adjusting for confounders.

**Results:** Analyses included 108,618 encounters (5,764 hospitalizations and 102,854 ED/UC encounters) among children and 309,483 encounters (76,072 hospitalizations and 233,411 ED/UC encounters) among adults. Among children across care settings, 17.0% (6,097/35,765) of cases versus 29.4% (21,449/72,853) of controls were vaccinated. Among adults, 28.2% (21,832/77,477) of cases versus 44.2% (102,560/232,006) of controls were vaccinated. VE was 51% (95% confidence interval [95% CI]: 41–60%) against influenza-associated hospitalizations and 54% (95% CI: 52–55%) against influenza-associated ED/UC encounters among children. VE was 43% (95% CI: 41–46%) against influenza-associated hospitalizations and 49% (95% CI: 47–50%) against influenza-associated ED/UC encounters among adults.

**Conclusions:** Influenza vaccination provided protection against influenza-associated hospitalizations and ED/UC encounters among children and adults in the US during the severe 2024–25 influenza season.

These findings support influenza vaccination as an important tool to reduce influenza-associated disease.

**Brief Summary:** During the 2024–25 influenza season, influenza vaccination provided protection against influenza-associated hospitalizations (43–51%) and emergency department or urgent care encounters (49–54%) among children and adults in the United States.

## INTRODUCTION

The 2024–25 influenza season was the first high severity season in the United States (US) since 2017–18, with high severity indicators observed in all age groups [1]. Over the course of the season, there were an estimated 23 million medical encounters, 710,000 hospitalizations, and 45,000 deaths due to influenza, with the highest hospitalization rates occurring among adults aged ≥65 years [1,2]. Influenza A viruses predominated, with co-circulation of A(H1N1)pdm09 and A(H3N2) viruses [3]. Although studies have shown that reference strains used for the 2024–25 Northern Hemisphere influenza vaccines were antigenically similar to circulating A(H1N1)pdm09 and B/Victoria viruses, some antigenic differences were identified between the 2024–25 vaccine strains and circulating A(H3N2) viruses [3].

Influenza vaccination was recommended for all persons aged ≥6 months in the US during the 2024–25 season [4]. Influenza vaccination has been shown to provide protection against a variety of influenza-associated outcomes, including symptomatic illness, outpatient visits, hospitalizations, and severe disease [5–8]. However, vaccine effectiveness (VE) may vary by season due to patient characteristics, antigenic similarity between vaccine and circulating influenza viruses, and other factors. Annual estimates from established VE platforms are important to assess the US influenza vaccination program [9].

We estimated the effectiveness of 2024–25 seasonal influenza vaccines against influenza-associated hospitalizations and emergency department (ED) or urgent care (UC) encounters among US children and adults.

## METHODS

### Population and Study Design

This analysis was conducted using data from the VIrtual SARS-CoV-2, Influenza, and Other respiratory viruses Network (VISION), a research collaboration between the US Centers for Disease Control and Prevention (CDC) and healthcare systems with integrated clinical, laboratory, and immunization records [10]. Detailed VISION methods have been published previously [11]. Briefly, participating healthcare systems capture electronic health record (EHR) data on medical encounters for acute respiratory illness (ARI) to evaluate the effectiveness of vaccines against influenza and other respiratory viruses using a test-negative, case-control design.

The current analysis included ARI-associated encounters occurring in the hospital and ED/UC settings from seven VISION sites in eight US states: Kaiser Permanente Northern California (California), Kaiser Permanente Southern California (California), University of Colorado (Colorado), Regenstrief Institute (Indiana), HealthPartners (Minnesota and Wisconsin), Kaiser Permanente Center for Health Research (Oregon and Washington), and Intermountain Health (Utah). ARI was defined using *International Classification of Diseases, Tenth Revision* (ICD-10) codes. Eligible encounters were those with ≥1 ARI-associated ICD-10 discharge diagnosis code and a molecular influenza test result among patients aged ≥6 months (Supplemental Table 1). Encounters were included from the date of the first influenza-positive case on or after October 1, 2024, by site and setting to the date of the last influenza-positive case on or before April 30, 2025, by site and setting (Supplemental Table 2). The same patient could contribute >1 encounter to the analysis. ED and/or UC encounters occurring within 7 days in the same patient were combined into a single ED/UC encounter. Hospitalizations occurring within 30 days in the same patient were combined into a single hospitalization.

Patient demographic and clinical characteristics were extracted from EHRs, including data on underlying medical conditions (ascertained using ICD-10 discharge diagnosis codes from the ED/UC or hospital encounter, Supplemental Table 3) and critical illness outcomes. Cases were defined as ARI-associated encounters with a positive molecular influenza test result from 10 days before to 72 hours after the encounter date. Controls were defined as ARI-associated encounters with a negative molecular influenza test result during the same window. Influenza vaccination status was determined from EHRs, state and local immunization information systems, and/or claims data. Data on vaccine product type were collected when available. Vaccinated encounters were defined as those with receipt of ≥1 dose of influenza vaccine on or after August 1, 2024 and ≥14 days before the index date (defined as the earlier date of either the most recent influenza test or the encounter date). Unvaccinated encounters had no documented receipt of influenza vaccine on or after August 1, 2024 and ≥14 days before the index date.

Encounters with missing or indeterminant results from a molecular influenza test, influenza vaccination <14 days before the index date, or a clinical diagnosis of influenza without a confirmatory test were excluded. Encounters with a positive molecular SARS-CoV-2 test result from 10 days before to 72 hours after the encounter date or a clinical diagnosis of COVID-19 were also excluded to reduce potential bias in vaccine effectiveness (VE) estimates due to correlation between influenza and COVID-19 vaccination behaviors [12,13].

### Statistical Analysis

Statistical analyses were conducted separately by age group and encounter setting (pediatric hospitalizations, pediatric ED/UC encounters, adult hospitalizations, adult ED/UC encounters). Demographic and clinical characteristics for each group were described by case and vaccination status, with a standardized mean difference of >0.20 between groups considered meaningful. VE was estimated using multivariable logistic regression comparing the odds of influenza vaccination among influenza-positive cases and influenza-negative controls. VE models were adjusted for age, sex, race and ethnicity, site, and calendar time. Age and calendar time were modeled as natural cubic splines with 4 degrees of freedom. VE was calculated as (1 – adjusted odds ratio) x 100 expressed as a percent.

VE estimates were stratified by time since vaccination (14–59, 60–119, ≥120 days), influenza type (A, B), and age group (6 months–4 years, 5–17 years, 18–49 years, 50–64 years, ≥65 years). Among pediatric and adult hospitalizations, VE estimates were also stratified by the presence of ≥1 immunocompromising condition (immunocompetent, immunocompromised), and VE was estimated against intensive care unit (ICU) admission and in-hospital death. VE against influenza-associated ICU admission was estimated by comparing the odds of influenza vaccination among influenza-positive cases who were admitted to an ICU and had no in-hospital death versus all influenza-negative hospitalized controls. VE against influenza-associated in-hospital death was estimated by comparing the odds of influenza vaccination among influenza-positive cases who had in-hospital death versus all influenza-negative hospitalized controls.

Among pediatric and adult ED/UC encounters, VE estimates were further stratified by setting (ED only, UC only).

Analyses were conducted in SAS version 9.4 (SAS Institute, Inc.) or R version 4.1.0 (R Foundation for Statistical Computing). This activity was reviewed by CDC and conducted consistent with applicable federal law and CDC policy (See e.g., 45C.F.R. part 46, 21C.F.R. part 56, 42 U.S.C. §241(d), 5 U.S.C. §552a, 44 U.S.C. §3501 et seq).

## RESULTS

### Included population

A total of 90,700 ARI-associated hospitalizations and 367,065 ARI-associated ED/UC encounters were identified. After excluding 8,864 (9.8%) hospitalizations and 30,800 (8.4%) ED/UC encounters (Supplemental Figure 1), 81,836 (90.2%) hospitalizations and 336,265 (91.6%) ED/UC encounters were included. Among 81,836 ARI-associated hospitalizations, 4,169 (5.1%) individuals had >1 encounter during the study period. Among 336,265 ARI-associated ED/UC encounters, 19,820 (5.9%) patients had >1 ED/UC encounter.

### Influenza circulation

Across sites, influenza activity peaked in early February (Supplemental Figures 2A-2D). Of 113,242 influenza cases, 101,489 (89.6%) were influenza A, 11,618 (10.3%) were influenza B, and 135 (0.1%) tested positive for >1 influenza virus. The proportion of influenza B cases was higher in children (15.9% of cases) than in adults (7.6% of cases).

**Figure 1.**
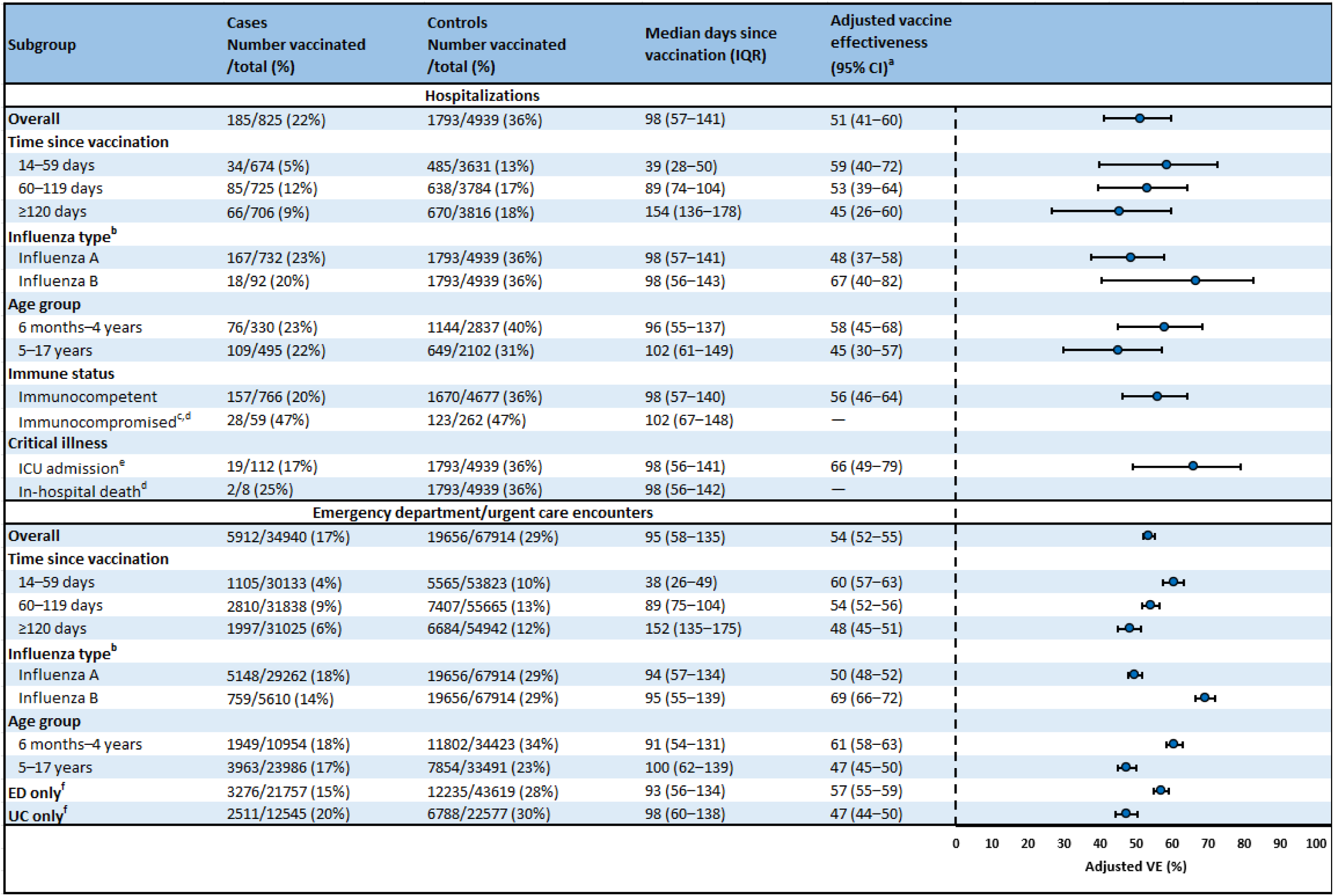
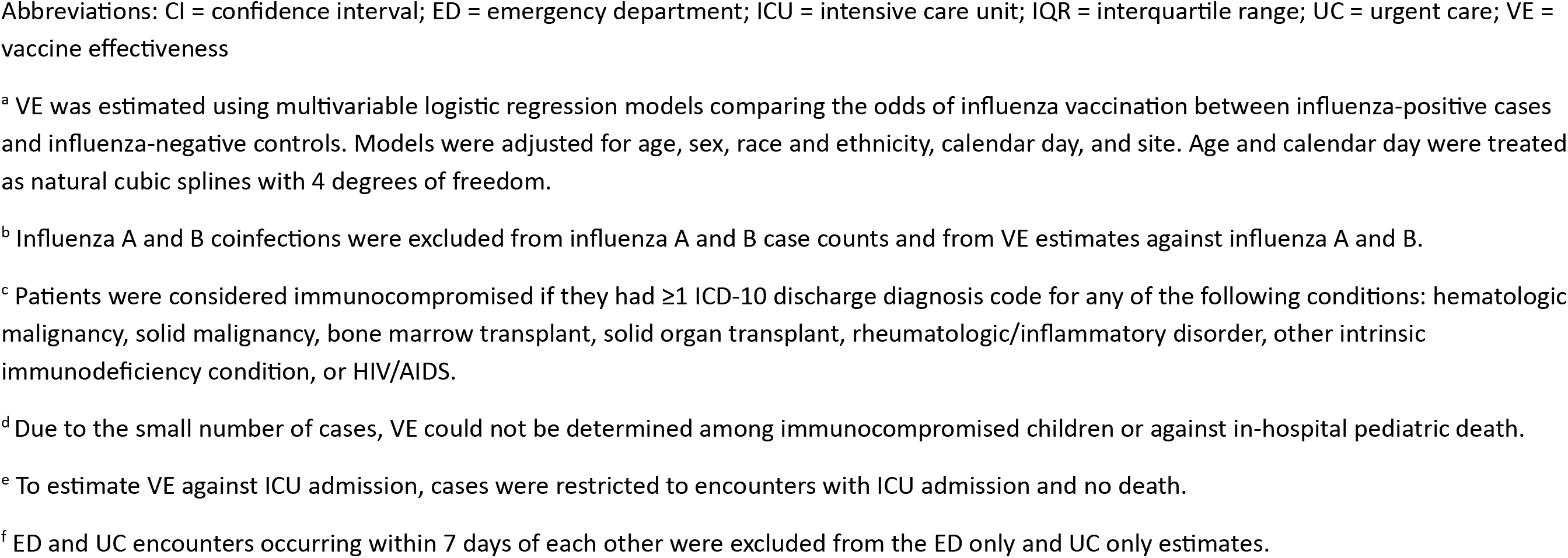
Influenza vaccine effectiveness against influenza-associated hospitalizations and emergency department or urgent care encounters among children aged 6 months–17 years — VISION, October 2024–April 2025

**Figure 2.**
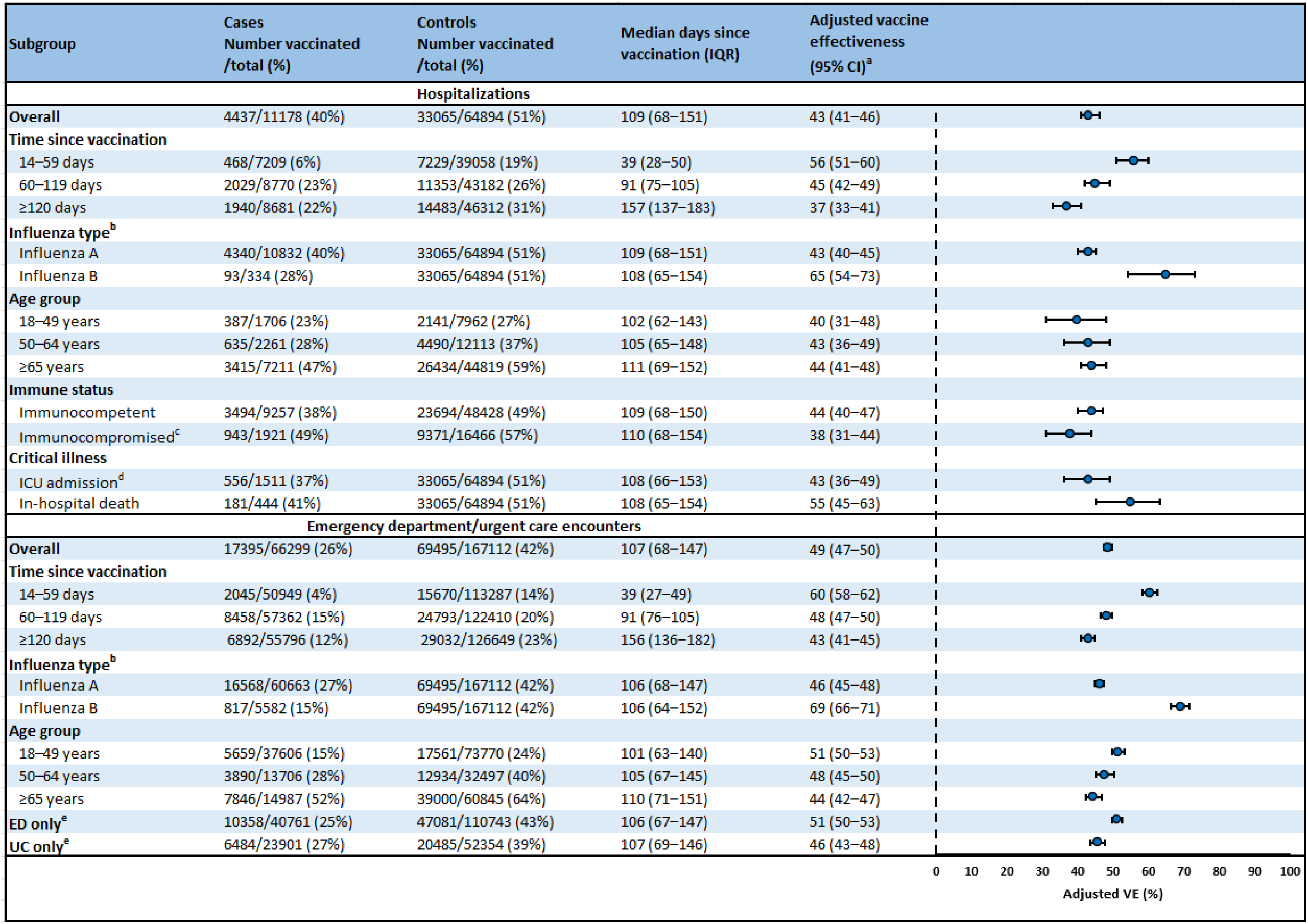
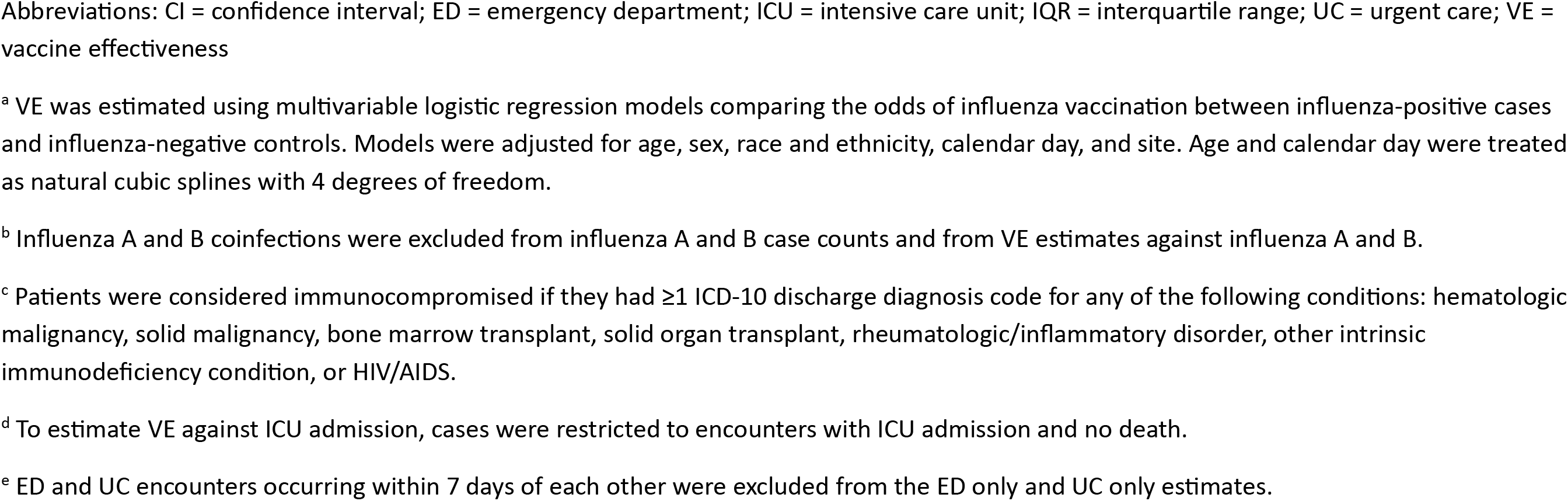
Influenza vaccine effectiveness against influenza-associated hospitalizations and emergency department or urgent care encounters among adults aged ≥18 years — VISION, October 2024–April 2025

### Pediatric patient characteristics

Of 5,764 ARI-associated hospitalizations in children, 825 (14.3%) were influenza-positive cases and 4,939 (85.7%) were influenza-negative controls (Table 1). Influenza positivity was 10.4% among children aged 6 months–4 years and 19.1% among those aged 5–17 years. Overall, 1,978 (34.3%) hospitalized children had received an influenza vaccine before their encounter (22.4% of cases versus 36.3% of controls, Figure 1). Among those vaccinated, median time since vaccination was 98 days. A total of 818 (14.2%) hospitalized children were admitted to an ICU during their admission and 30 (0.5%) died.

**Table 1.** Characteristics of acute respiratory illness-associated hospitalizations and emergency department/urgent care encounters among children aged 6 months–17 years by influenza vaccination and test status — VISION, October 2024–April 2025

|  |  | Influenza vaccination status |  |  | Influenza test result |  |  |
| --- | --- | --- | --- | --- | --- | --- | --- |
|  | Total<br>(col %) | Vaccinated<br>(row %) | Unvaccinated<br>(row %) | SMD<sup>a</sup> | Positive<br>(row %) | Negative<br>(row %) | SMD<sup>a</sup> |
| **All hospitalizations** | 5764 | 1978 (34.3) | 3786 (65.7) |  | 825 (14.3) | 4939 (85.7) |  |
| Influenza vaccination status |  |  |  |  |  |  |  |
| Unvaccinated | 3786 (65.7) | 0 (0.0) | 3786 (100.0) | -- | 640 (16.9) | 3146 (83.1) | 0.308 |
| Vaccinated | 1978 (34.3) | 1978 (100.0) | 0 (0.0) |  | 185 (9.4) | 1793 (90.6) |  |
| 14–59 days earlier | 519 (9.0) | 519 (100.0) | 0 (0.0) |  | 34 (6.6) | 485 (93.4) |  |
| 60–119 days earlier | 723 (12.5) | 723 (100.0) | 0 (0.0) |  | 85 (11.8) | 638 (88.2) |  |
| ≥120 days earlier | 736 (12.8) | 736 (100.0) | 0 (0.0) |  | 66 (9.0) | 670 (91.0) |  |
| Month of encounter |  |  |  |  |  |  |  |
| October 2024 | 346 (6.0) | 63 (18.2) | 283 (81.8) | 0.289 | 4 (1.2) | 342 (98.8) | 0.745 |
| November 2024 | 637 (11.1) | 165 (25.9) | 472 (74.1) |  | 24 (3.8) | 613 (96.2) |  |
| December 2024 | 962 (16.7) | 295 (30.7) | 667 (69.3) |  | 169 (17.6) | 793 (82.4) |  |
| January 2025 | 991 (17.2) | 339 (34.2) | 652 (65.8) |  | 242 (24.4) | 749 (75.6) |  |
| February 2025 | 1193 (20.7) | 450 (37.7) | 743 (62.3) |  | 253 (21.2) | 940 (78.8) |  |
| March 2025 | 955 (16.6) | 392 (41.0) | 563 (59.0) |  | 94 (9.8) | 861 (90.2) |  |
| April 2025 | 680 (11.8) | 274 (40.3) | 406 (59.7) |  | 39 (5.7) | 641 (94.3) |  |
| Site |  |  |  |  |  |  |  |
| Site A | 113 (2.0) | 39 (34.5) | 74 (65.5) | 0.216 | 8 (7.1) | 105 (92.9) | 0.180 |
| Site B | 21 (0.4) | 6 (28.6) | 15 (71.4) |  | 3 (14.3) | 18 (85.7) |  |
| Site C | 2133 (37.0) | 703 (33.0) | 1430 (67.0) |  | 264 (12.4) | 1869 (87.6) |  |
| Site D | 257 (4.5) | 86 (33.5) | 171 (66.5) |  | 35 (13.6) | 222 (86.4) |  |
| Site E | 901 (15.6) | 229 (25.4) | 672 (74.6) |  | 137 (15.2) | 764 (84.8) |  |
| Site F | 1085 (18.8) | 415 (38.2) | 670 (61.8) |  | 191 (17.6) | 894 (82.4) |  |
| Site G | 1254 (21.8) | 500 (39.9) | 754 (60.1) |  | 187 (14.9) | 1067 (85.1) |  |
| Median age (IQR), years | 4 (1.8, 8) | 3 (1.4, 7) | 4 (2, 9) |  | 6 (2, 10) | 3 (1.6, 8) |  |
| Age group |  |  |  |  |  |  |  |
| 6 months–4 years | 3167 (54.9) | 1220 (38.5) | 1947 (61.5) | 0.208 | 330 (10.4) | 2837 (89.6) | 0.354 |
| 5–17 years | 2597 (45.1) | 758 (29.2) | 1839 (70.8) |  | 495 (19.1) | 2102 (80.9) |  |
| Female | 2596 (45.0) | 883 (34.0) | 1713 (66.0) | 0.012 | 378 (14.6) | 2218 (85.4) | 0.018 |
| Race/ethnicity |  |  |  |  |  |  |  |
| White Non-Hispanic | 2634 (45.7) | 871 (33.1) | 1763 (66.9) | 0.196 | 336 (12.8) | 2298 (87.2) | 0.135 |
| Black Non-Hispanic | 425 (7.4) | 118 (27.8) | 307 (72.2) |  | 78 (18.4) | 347 (81.6) |  |
| Other Non-Hispanic<sup>b</sup> | 710 (12.3) | 325 (45.8) | 385 (54.2) |  | 102 (14.4) | 608 (85.6) |  |
| Hispanic | 1689 (29.3) | 564 (33.4) | 1125 (66.6) |  | 264 (15.6) | 1425 (84.4) |  |
| Unknown | 306 (5.3) | 100 (32.7) | 206 (67.3) |  | 45 (14.7) | 261 (85.3) |  |
| Any medical condition<sup>c</sup> | 3625 (62.9) | 1336 (36.9) | 2289 (63.1) | 0.148 | 491 (13.5) | 3134 (86.5) | 0.081 |
| Respiratory condition | 2666 (46.3) | 996 (37.4) | 1670 (62.6) | 0.125 | 298 (11.2) | 2368 (88.8) | 0.241 |
| Non-respiratory condition | 1829 (31.7) | 735 (40.2) | 1094 (59.8) | 0.176 | 308 (16.8) | 1521 (83.2) | 0.138 |
| Immunocompromising condition<sup>d</sup> | 321 (5.6) | 151 (47.0) | 170 (53.0) | 0.132 | 59 (18.4) | 262 (81.6) | 0.076 |
| Influenza type |  |  |  |  |  |  |  |
| Influenza A only | 732 (88.7) | 167 (22.8) | 565 (77.2) | 0.082 | 732 (100.0) | 0 (0.0) | -- |
| Influenza B only | 92 (11.2) | 18 (19.6) | 74 (80.4) |  | 92 (100.0) | 0 (0.0) |  |
| Influenza A and B | 1 (0.1) | 0 (0.0) | 1 (100.0) |  | 1 (100.0) | 0 (0.0) |  |
| ICU admission |  |  |  |  |  |  |  |
| Yes | 818 (14.2) | 302 (36.9) | 516 (63.1) | 0.048 | 120 (14.7) | 698 (85.3) | 0.032 |
| No | 4866 (84.4) | 1647 (33.8) | 3219 (66.2) |  | 691 (14.2) | 4175 (85.8) |  |
| Unknown | 80 (1.4) | 29 (36.2) | 51 (63.7) |  | 14 (17.5) | 66 (82.5) |  |
| In-hospital death |  |  |  |  |  |  |  |
| Yes | 30 (0.5) | 9 (30.0) | 21 (70.0) | 0.014 | 8 (26.7) | 22 (73.3) | 0.063 |
| No | 5734 (99.5) | 1969 (34.3) | 3765 (65.7) |  | 817 (14.2) | 4917 (85.8) |  |
| Unknown | 0 (0.0) | 0 (0.0) | 0 (0.0) |  | 0 (0.0) | 0 (0.0) |  |
| **All ED/UC encounters** | 102854 | 25568 (24.9) | 77286 (75.1) |  | 34940 (34.0) | 67914 (66.0) |  |
| Influenza vaccination status |  |  |  |  |  |  |  |
| Unvaccinated | 77286 (75.1) | 0 (0.0) | 77286 (100.0) | -- | 29028 (37.6) | 48258 (62.4) | 0.289 |
| Vaccinated | 25568 (24.9) | 25568 (100.0) | 0 (0.0) |  | 5912 (23.1) | 19656 (76.9) |  |
| 14–59 days earlier | 6670 (6.5) | 6670 (100.0) | 0 (0.0) |  | 1105 (16.6) | 5565 (83.4) |  |
| 60–119 days earlier | 10217 (9.9) | 10217 (100.0) | 0 (0.0) |  | 2810 (27.5) | 7407 (72.5) |  |
| ≥120 days earlier | 8681 (8.4) | 8681 (100.0) | 0 (0.0) |  | 1997 (23.0) | 6684 (77.0) |  |
| Month of encounter |  |  |  |  |  |  |  |
| October 2024 | 7594 (7.4) | 716 (9.4) | 6878 (90.6) | 0.341 | 174 (2.3) | 7420 (97.7) | 0.841 |
| November 2024 | 10017 (9.7) | 1984 (19.8) | 8033 (80.2) |  | 988 (9.9) | 9029 (90.1) |  |
| December 2024 | 20065 (19.5) | 4482 (22.3) | 15583 (77.7) |  | 7996 (39.9) | 12069 (60.1) |  |
| January 2025 | 20903 (20.3) | 5098 (24.4) | 15805 (75.6) |  | 10592 (50.7) | 10311 (49.3) |  |
| February 2025 | 20896 (20.3) | 5711 (27.3) | 15185 (72.7) |  | 9992 (47.8) | 10904 (52.2) |  |
| March 2025 | 14127 (13.7) | 4409 (31.2) | 9718 (68.8) |  | 3841 (27.2) | 10286 (72.8) |  |
| April 2025 | 9252 (9.0) | 3168 (34.2) | 6084 (65.8) |  | 1357 (14.7) | 7895 (85.3) |  |
| Site |  |  |  |  |  |  |  |
| Site A | 2695 (2.6) | 654 (24.3) | 2041 (75.7) | 0.251 | 1023 (38.0) | 1672 (62.0) | 0.164 |
| Site B | 5756 (5.6) | 1498 (26.0) | 4258 (74.0) |  | 2120 (36.8) | 3636 (63.2) |  |
| Site C | 16049 (15.6) | 3939 (24.5) | 12110 (75.5) |  | 4879 (30.4) | 11170 (69.6) |  |
| Site D | 14009 (13.6) | 3598 (25.7) | 10411 (74.3) |  | 3971 (28.3) | 10038 (71.7) |  |
| Site E | 14698 (14.3) | 2132 (14.5) | 12566 (85.5) |  | 4867 (33.1) | 9831 (66.9) |  |
| Site F | 21465 (20.9) | 5912 (27.5) | 15553 (72.5) |  | 7179 (33.4) | 14286 (66.6) |  |
| Site G | 28182 (27.4) | 7835 (27.8) | 20347 (72.2) |  | 10901 (38.7) | 17281 (61.3) |  |
| Median age (IQR), years | 5 (2, 10) | 4 (1.7, 9) | 6 (2, 11) |  | 7 (4, 11) | 4 (2, 10) |  |
| Age group |  |  |  |  |  |  |  |
| 6 months–4 years | 45377 (44.1) | 13751 (30.3) | 31626 (69.7) | 0.260 | 10954 (24.1) | 34423 (75.9) | 0.401 |
| 5–17 years | 57477 (55.9) | 11817 (20.6) | 45660 (79.4) |  | 23986 (41.7) | 33491 (58.3) |  |
| Female | 47676 (46.4) | 11530 (24.2) | 36146 (75.8) | 0.034 | 16671 (35.0) | 31005 (65.0) | 0.041 |
| Race/ethnicity |  |  |  |  |  |  |  |
| White Non-Hispanic | 39537 (38.4) | 9394 (23.8) | 30143 (76.2) | 0.249 | 12206 (30.9) | 27331 (69.1) | 0.112 |
| Black Non-Hispanic | 10078 (9.8) | 1613 (16.0) | 8465 (84.0) |  | 3703 (36.7) | 6375 (63.3) |  |
| Other Non-Hispanic<sup>b</sup> | 13184 (12.8) | 4641 (35.2) | 8543 (64.8) |  | 4644 (35.2) | 8540 (64.8) |  |
| Hispanic | 35310 (34.3) | 8770 (24.8) | 26540 (75.2) |  | 12756 (36.1) | 22554 (63.9) |  |
| Unknown | 4745 (4.6) | 1150 (24.2) | 3595 (75.8) |  | 1631 (34.4) | 3114 (65.6) |  |
| Influenza type |  |  |  |  |  |  |  |
| Influenza A only | 29262 (83.7) | 5148 (17.6) | 24114 (82.4) | 0.115 | 29262 (100.0) | 0 (0.0) | -- |
| Influenza B only | 5610 (16.1) | 759 (13.5) | 4851 (86.5) |  | 5610 (100.0) | 0 (0.0) |  |
| Influenza A and B | 68 (0.2) | 5 (7.4) | 63 (92.6) |  | 68 (100.0) | 0 (0.0) |  |

Abbreviations: ED/UC = emergency department/urgent care; ICU = intensive care unit; IQR = interquartile range; SMD = standardized mean difference
<sup>a</sup> An absolute SMD >0.20 indicates a non-negligible difference in variable distributions between vaccinated versus unvaccinated encounters or for encounters with positive influenza test results versus negative influenza test results.
<sup>b</sup> Other race is defined as any one of the following responses: Asian, Hawaiian or other Pacific Islander, American Indian or Alaska Native, Middle Eastern or North African, Other, or multiple races.
<sup>c</sup> Includes respiratory conditions (asthma, COPD, other lung conditions) and non-respiratory conditions (cardiovascular, neurologic, hematologic, endocrine, renal, gastrointestinal).
<sup>d</sup> Patients were considered immunocompromised if they had ≥1 ICD-10 discharge diagnosis code for any of the following conditions: hematologic malignancy, solid malignancy, bone marrow transplant, solid organ transplant, rheumatologic/inflammatory disorder, other intrinsic immunodeficiency condition, or HIV/AIDS.

Of 102,854 ARI-associated ED/UC encounters in children, 34,940 (34.0%) were influenza-positive cases and 67,914 (66.0%) were influenza-negative controls (Table 1). Influenza positivity was 24.1% among children aged 6 months–4 years and 41.7% among those aged 5–17 years. Overall, 25,568 (24.9%) had received an influenza vaccine before the encounter (16.9% of cases versus 28.9% of controls, Figure 1). Among vaccinated patients, median time since vaccination was 95 days.

Across care settings, 84.5% of vaccinated children with a known product type received standard-dose, egg-based inactivated vaccine, 9.8% received cell-culture-based vaccine, 4.0% received live-attenuated vaccine, and 1.7% received a different product type (Supplemental Table 4).

### Pediatric vaccine effectiveness

Overall VE against pediatric influenza-associated hospitalizations was 51% (95% confidence interval [95% CI]: 41–60) (Figure 1). VE among children aged 6 months–4 years was 58% (95% CI: 45-68), and VE among children aged 5–17 years was 45% (95% CI: 30–57). VE was 48% (95% CI: 37–58) against influenza A and 67% (95% CI: 40–82) against influenza B. VE against ICU admission was 66% (95% CI: 49–79). Due to the small number of cases, VE could not be determined among immunocompromised children or against in-hospital pediatric death.

Overall VE against pediatric influenza-associated ED/UC encounters was 54% (95% CI: 52–55%) (Figure 1). VE among children aged 6 months–4 years was 61% (95% CI: 58–63), and VE among children aged 5–17 years was 47% (95% CI: 45–50). VE was 50% (95% CI: 48–52) against influenza A and 69% (95% CI: 66–72) against influenza B.

### Adult patient characteristics

Of 76,072 ARI-associated hospitalizations in adults, 11,178 (14.7%) were influenza-positive cases and 64,894 (85.3%) were influenza-negative controls (Table 2). Influenza positivity was 17.6% among adults aged 18–49 years, 15.7% among adults aged 50–64 years, and 13.9% among adults aged ≥65 years.

**Table 2.** Characteristics of acute respiratory illness-associated hospitalizations and emergency department/urgent care encounters among adults aged ≥18 years by influenza vaccination and test status — VISION, October 2024–April 2025

|  |  | Influenza vaccination status |  |  | Influenza test result |  |  |
| --- | --- | --- | --- | --- | --- | --- | --- |
|  | Total<br>(col %) | Vaccinated<br>(row %) | Unvaccinated<br>(row %) | SMD<sup>a</sup> | Positive<br>(row %) | Negative<br>(row %) | SMD<sup>a</sup> |
| **All hospitalizations** | 76072 | 37502 (49.3) | 38570 (50.7) |  | 11178 (14.7) | 64894 (85.3) |  |
| Influenza vaccination status |  |  |  |  |  |  |  |
| Unvaccinated | 38570 (50.7) | 0 (0.0) | 38570 (100.0) | -- | 6741 (17.5) | 31829 (82.5) | 0.228 |
| Vaccinated | 37502 (49.3) | 37502 (100.0) | 0 (0.0) |  | 4437 (11.8) | 33065 (88.2) |  |
| 14–59 days earlier | 7697 (10.1) | 7697 (100.0) | 0 (0.0) |  | 468 (6.1) | 7229 (93.9) |  |
| 60–119 days earlier | 13382 (17.6) | 13382 (100.0) | 0 (0.0) |  | 2029 (15.2) | 11353 (84.8) |  |
| ≥120 days earlier | 16423 (21.6) | 16423 (100.0) | 0 (0.0) |  | 1940 (11.8) | 14483 (88.2) |  |
| Month of encounter |  |  |  |  |  |  |  |
| October 2024 | 5386 (7.1) | 1602 (29.7) | 3784 (70.3) | 0.273 | 50 (0.9) | 5336 (99.1) | 0.919 |
| November 2024 | 8311 (10.9) | 3559 (42.8) | 4752 (57.2) |  | 225 (2.7) | 8086 (97.3) |  |
| December 2024 | 12453 (16.4) | 5912 (47.5) | 6541 (52.5) |  | 2149 (17.3) | 10304 (82.7) |  |
| January 2025 | 14921 (19.6) | 7493 (50.2) | 7428 (49.8) |  | 3729 (25.0) | 11192 (75.0) |  |
| February 2025 | 13468 (17.7) | 6934 (51.5) | 6534 (48.5) |  | 3526 (26.2) | 9942 (73.8) |  |
| March 2025 | 12102 (15.9) | 6569 (54.3) | 5533 (45.7) |  | 1213 (10.0) | 10889 (90.0) |  |
| April 2025 | 9431 (12.4) | 5433 (57.6) | 3998 (42.4) |  | 286 (3.0) | 9145 (97.0) |  |
| Site |  |  |  |  |  |  |  |
| Site A | 2741 (3.6) | 1339 (48.9) | 1402 (51.1) | 0.482 | 413 (15.1) | 2328 (84.9) | 0.097 |
| Site B | 3783 (5.0) | 2041 (54.0) | 1742 (46.0) |  | 630 (16.7) | 3153 (83.3) |  |
| Site C | 7668 (10.1) | 2884 (37.6) | 4784 (62.4) |  | 1117 (14.6) | 6551 (85.4) |  |
| Site D | 9322 (12.3) | 3698 (39.7) | 5624 (60.3) |  | 1171 (12.6) | 8151 (87.4) |  |
| Site E | 19090 (25.1) | 6949 (36.4) | 12141 (63.6) |  | 3101 (16.2) | 15989 (83.8) |  |
| Site F | 20401 (26.8) | 12094 (59.3) | 8307 (40.7) |  | 2890 (14.2) | 17511 (85.8) |  |
| Site G | 13067 (17.2) | 8497 (65.0) | 4570 (35.0) |  | 1856 (14.2) | 11211 (85.8) |  |
| Median age (IQR), years | 72 (61, 81) | 76 (67, 84) | 68 (55, 78) |  | 71 (59, 80) | 72 (61, 82) |  |
| Age group |  |  |  |  |  |  |  |
| 18–49 years | 9668 (12.7) | 2528 (26.1) | 7140 (73.9) | 0.502 | 1706 (17.6) | 7962 (82.4) | 0.104 |
| 50–64 years | 14374 (18.9) | 5125 (35.7) | 9249 (64.3) |  | 2261 (15.7) | 12113 (84.3) |  |
| ≥65 years | 52030 (68.4) | 29849 (57.4) | 22181 (42.6) |  | 7211 (13.9) | 44819 (86.1) |  |
| Female | 40280 (52.9) | 19921 (49.5) | 20359 (50.5) | 0.007 | 6153 (15.3) | 34127 (84.7) | 0.049 |
| Race/ethnicity |  |  |  |  |  |  |  |
| White Non-Hispanic | 49902 (65.6) | 24496 (49.1) | 25406 (50.9) | 0.191 | 7228 (14.5) | 42674 (85.5) | 0.034 |
| Black Non-Hispanic | 7482 (9.8) | 3000 (40.1) | 4482 (59.9) |  | 1095 (14.6) | 6387 (85.4) |  |
| Other Non-Hispanic<sup>b</sup> | 7334 (9.6) | 4294 (58.5) | 3040 (41.5) |  | 1123 (15.3) | 6211 (84.7) |  |
| Hispanic | 10243 (13.5) | 5362 (52.3) | 4881 (47.7) |  | 1586 (15.5) | 8657 (84.5) |  |
| Unknown | 1111 (1.5) | 350 (31.5) | 761 (68.5) |  | 146 (13.1) | 965 (86.9) |  |
| Any medical condition<sup>c</sup> | 70047 (92.1) | 35643 (50.9) | 34404 (49.1) | 0.218 | 9722 (13.9) | 60325 (86.1) | 0.200 |
| Respiratory condition | 37568 (49.4) | 19909 (53.0) | 17659 (47.0) | 0.146 | 4817 (12.8) | 32751 (87.2) | 0.148 |
| Non-respiratory condition | 66663 (87.6) | 34530 (51.8) | 32133 (48.2) | 0.269 | 9110 (13.7) | 57553 (86.3) | 0.203 |
| Immunocompromising condition<sup>d</sup> | 18387 (24.2) | 10314 (56.1) | 8073 (43.9) | 0.154 | 1921 (10.4) | 16466 (89.6) | 0.201 |
| Influenza type |  |  |  |  |  |  |  |
| Influenza A only | 10832 (96.9) | 4340 (40.1) | 6492 (59.9) | 0.090 | 10832 (100.0) | 0 (0.0) | -- |
| Influenza B only | 334 (3.0) | 93 (27.8) | 241 (72.2) |  | 334 (100.0) | 0 (0.0) |  |
| Influenza A and B | 12 (0.1) | 4 (33.3) | 8 (66.7) |  | 12 (100.0) | 0 (0.0) |  |
| ICU admission |  |  |  |  |  |  |  |
| Yes | 15456 (20.3) | 7389 (47.8) | 8067 (52.2) | 0.039 | 1755 (11.4) | 13701 (88.6) | 0.141 |
| No | 60249 (79.2) | 29898 (49.6) | 30351 (50.4) |  | 9379 (15.6) | 50870 (84.4) |  |
| Unknown | 367 (0.5) | 215 (58.6) | 152 (41.4) |  | 44 (12.0) | 323 (88.0) |  |
| In-hospital death |  |  |  |  |  |  |  |
| Yes | 5335 (7.0) | 2813 (52.7) | 2522 (47.3) | 0.038 | 444 (8.3) | 4891 (91.7) | 0.154 |
| No | 70736 (93.0) | 34689 (49.0) | 36047 (51.0) |  | 10734 (15.2) | 60002 (84.8) |  |
| Unknown | 1 (0.0) | 0 (0.0) | 1 (100.0) |  | 0 (0.0) | 1 (100.0) |  |
| **All ED/UC encounters** | 233411 | 86890 (37.2) | 146521 (62.8) |  | 66299 (28.4) | 167112 (71.6) |  |
| Influenza vaccination status |  |  |  |  |  |  |  |
| Unvaccinated | 146521 (62.8) | 0 (0.0) | 146521 (100.0) | -- | 48904 (33.4) | 97617 (66.6) | 0.329 |
| Vaccinated | 86890 (37.2) | 86890 (100.0) | 0 (0.0) |  | 17395 (20.0) | 69495 (80.0) |  |
| 14–59 days earlier | 17715 (7.6) | 17715 (100.0) | 0 (0.0) |  | 2045 (11.5) | 15670 (88.5) |  |
| 60–119 days earlier | 33251 (14.2) | 33251 (100.0) | 0 (0.0) |  | 8458 (25.4) | 24793 (74.6) |  |
| ≥120 days earlier | 35924 (15.4) | 35924 (100.0) | 0 (0.0) |  | 6892 (19.2) | 29032 (80.8) |  |
| Month of encounter |  |  |  |  |  |  |  |
| October 2024 | 17282 (7.4) | 3619 (20.9) | 13663 (79.1) | 0.271 | 460 (2.7) | 16822 (97.3) | 0.848 |
| November 2024 | 22232 (9.5) | 7645 (34.4) | 14587 (65.6) |  | 1902 (8.6) | 20330 (91.4) |  |
| December 2024 | 45178 (19.4) | 15469 (34.2) | 29709 (65.8) |  | 16149 (35.7) | 29029 (64.3) |  |
| January 2025 | 52760 (22.6) | 19442 (36.8) | 33318 (63.2) |  | 23341 (44.2) | 29419 (55.8) |  |
| February 2025 | 41490 (17.8) | 16061 (38.7) | 25429 (61.3) |  | 16326 (39.3) | 25164 (60.7) |  |
| March 2025 | 31174 (13.4) | 13672 (43.9) | 17502 (56.1) |  | 6030 (19.3) | 25144 (80.7) |  |
| April 2025 | 23295 (10.0) | 10982 (47.1) | 12313 (52.9) |  | 2091 (9.0) | 21204 (91.0) |  |
| Site |  |  |  |  |  |  |  |
| Site A | 10075 (4.3) | 4003 (39.7) | 6072 (60.3) | 0.382 | 2744 (27.2) | 7331 (72.8) | 0.137 |
| Site B | 16024 (6.9) | 6007 (37.5) | 10017 (62.5) |  | 4315 (26.9) | 11709 (73.1) |  |
| Site C | 28168 (12.1) | 7450 (26.4) | 20718 (73.6) |  | 8234 (29.2) | 19934 (70.8) |  |
| Site D | 26121 (11.2) | 8309 (31.8) | 17812 (68.2) |  | 6288 (24.1) | 19833 (75.9) |  |
| Site E | 28977 (12.4) | 6271 (21.6) | 22706 (78.4) |  | 7546 (26.0) | 21431 (74.0) |  |
| Site F | 55862 (23.9) | 24084 (43.1) | 31778 (56.9) |  | 15181 (27.2) | 40681 (72.8) |  |
| Site G | 68184 (29.2) | 30766 (45.1) | 37418 (54.9) |  | 21991 (32.3) | 46193 (67.7) |  |
| Median age (IQR), years | 51 (34, 70) | 67 (48, 78) | 43 (30, 60) |  | 45 (31, 63) | 54 (35, 72) |  |
| Age group |  |  |  |  |  |  |  |
| 18–49 years | 111376 (47.7) | 23220 (20.8) | 88156 (79.2) | 0.818 | 37606 (33.8) | 73770 (66.2) | 0.315 |
| 50–64 years | 46203 (19.8) | 16824 (36.4) | 29379 (63.6) |  | 13706 (29.7) | 32497 (70.3) |  |
| ≥65 years | 75832 (32.5) | 46846 (61.8) | 28986 (38.2) |  | 14987 (19.8) | 60845 (80.2) |  |
| Female | 138796 (59.5) | 52572 (37.9) | 86224 (62.1) | 0.034 | 38534 (27.8) | 100262 (72.2) | 0.038 |
| Race/ethnicity |  |  |  |  |  |  |  |
| White Non-Hispanic | 118813 (50.9) | 45799 (38.5) | 73014 (61.5) | 0.202 | 29755 (25.0) | 89058 (75.0) | 0.197 |
| Black Non-Hispanic | 23755 (10.2) | 6954 (29.3) | 16801 (70.7) |  | 6532 (27.5) | 17223 (72.5) |  |
| Other Non-Hispanic<sup>b</sup> | 27021 (11.6) | 12453 (46.1) | 14568 (53.9) |  | 7994 (29.6) | 19027 (70.4) |  |
| Hispanic | 58249 (25.0) | 20333 (34.9) | 37916 (65.1) |  | 19953 (34.3) | 38296 (65.7) |  |
| Unknown | 5573 (2.4) | 1351 (24.2) | 4222 (75.8) |  | 2065 (37.1) | 3508 (62.9) |  |
| Influenza type |  |  |  |  |  |  |  |
| Influenza A only | 60663 (91.5) | 16568 (27.3) | 44095 (72.7) | 0.196 | 60663 (100.0) | 0 (0.0) | -- |
| Influenza B only | 5582 (8.4) | 817 (14.6) | 4765 (85.4) |  | 5582 (100.0) | 0 (0.0) |  |
| Influenza A and B | 54 (0.1) | 10 (18.5) | 44 (81.5) |  | 54 (100.0) | 0 (0.0) |  |

Abbreviations: ED/UC = emergency department/urgent care; ICU = intensive care unit; IQR = interquartile range; SMD = standardized mean difference
<sup>a</sup> An absolute SMD >0.20 indicates a non-negligible difference in variable distributions between vaccinated versus unvaccinated encounters or for encounters with positive influenza test results versus negative influenza test results.
<sup>b</sup> Other race is defined as any one of the following responses: Asian, Hawaiian or other Pacific Islander, American Indian or Alaska Native, Other, Middle Eastern or North African, or multiple races.
<sup>c</sup> Includes respiratory conditions (asthma, COPD, other lung conditions) and non-respiratory conditions (cardiovascular, neurologic, hematologic, endocrine, renal, gastrointestinal).
<sup>d</sup> Patients were considered immunocompromised if they had ≥1 ICD-10 discharge diagnosis code for any of the following conditions: hematologic malignancy, solid malignancy, bone marrow transplant, solid organ transplant, rheumatologic/inflammatory disorder, other intrinsic immunodeficiency condition, or HIV/AIDS.

Median age was 71 years for cases and 72 years for controls. Overall, 37,502 (49.3%) hospitalized adults had received an influenza vaccine before their encounter (39.7% of cases versus 51.0% of controls, Figure 2). Among those vaccinated, median time since vaccination was 109 days. A total of 15,456 (20.3%) hospitalized adults were admitted to an ICU during their admission and 5,335 (7.0%) died.

Of 233,411 ARI-associated ED/UC encounters in adults, 66,299 (28.4%) were influenza-positive cases and 167,112 (71.6%) were influenza-negative controls (Table 2). Influenza positivity was 33.8% among adults aged 18–49 years, 29.7% among adults aged 50–64 years, and 19.8% among adults aged ≥65 years.

Median age was 45 years for cases and 54 years for controls. Overall, 86,890 (37.2%) had received an influenza vaccine before the encounter (26.2% of cases versus 41.6% of controls, Figure 2). Among vaccinated patients, median time since vaccination was 107 days.

Across care settings, 83.7% of vaccinated adults aged 18–64 years with a known product type received a standard-dose, egg-based inactivated vaccine (Supplemental Table 4). Among adults aged ≥65 years, 92.4% received a high-dose inactivated, adjuvanted, or recombinant vaccine, all of which were preferentially recommended for adults aged ≥65 years [4].

### Adult vaccine effectiveness

Overall VE against influenza-associated hospitalizations was 43% (95% CI: 41–46) among adults (Figure 2). When stratified by age group, VE was 40% (95% CI: 31–48) among adults aged 18–49 years, 43% (95% CI: 36–49) among adults aged 50–64 years, and 44% (95% CI: 41–48) among adults aged ≥65 years. VE was 43% (95% CI: 40–45) against influenza A and 65% (95% CI: 54–73) against influenza B. Among immunocompetent and immunocompromised adults, VE estimates were 44% (95% CI: 40–47) and 38% (95% CI: 31–44), respectively. VE was 43% (95% CI: 36–49) against ICU admission and 55% (95% CI: 45–63) against in-hospital death.

Overall VE against influenza-associated ED/UC encounters was 49% (95% CI: 47–50) among adults (Figure 2). When stratified by age group, VE was 51% (95% CI: 50–53) among adults aged 18–49 years, 48% (95% CI: 45–50) among adults aged 50–64 years, and 44% (95% CI: 42–47) among adults aged ≥65 years. VE was 46% (95% CI: 45–48) against influenza A and 69% (95% CI: 66–71) against influenza B.

### Vaccine effectiveness by time since vaccination

Among both children and adults, influenza vaccination provided protection against influenza-associated hospitalizations and ED/UC encounters through ≥120 days after vaccination (Figures 1-3, Supplemental Table 5). Among children, VE against hospitalizations was 59% (95% CI: 40–72) at 14–59 days, 53% (95% CI: 39–64) at 60–119 days, and 45% (95% CI: 26–60) at ≥120 days after vaccination (Figure 1). VE against ED/UC encounters was 60% (95% CI: 57–63) at 14–59 days, 54% (95% CI: 52–56) at 60–119 days, and 48% (95% CI: 45–51) at ≥120 days after vaccination. VE estimates stratified by influenza type, age group, and time since vaccination are shown in Figure 3 and Supplemental Table 5. Among children aged 6 months–4 years, VE did not decline against influenza A or influenza B-associated ED/UC encounters through ≥120 days after vaccination. However, among children aged 5–17 years, VE estimates against influenza A and influenza B were lower at ≥120 days compared to 14–59 days after vaccination.

**Figure 3.**
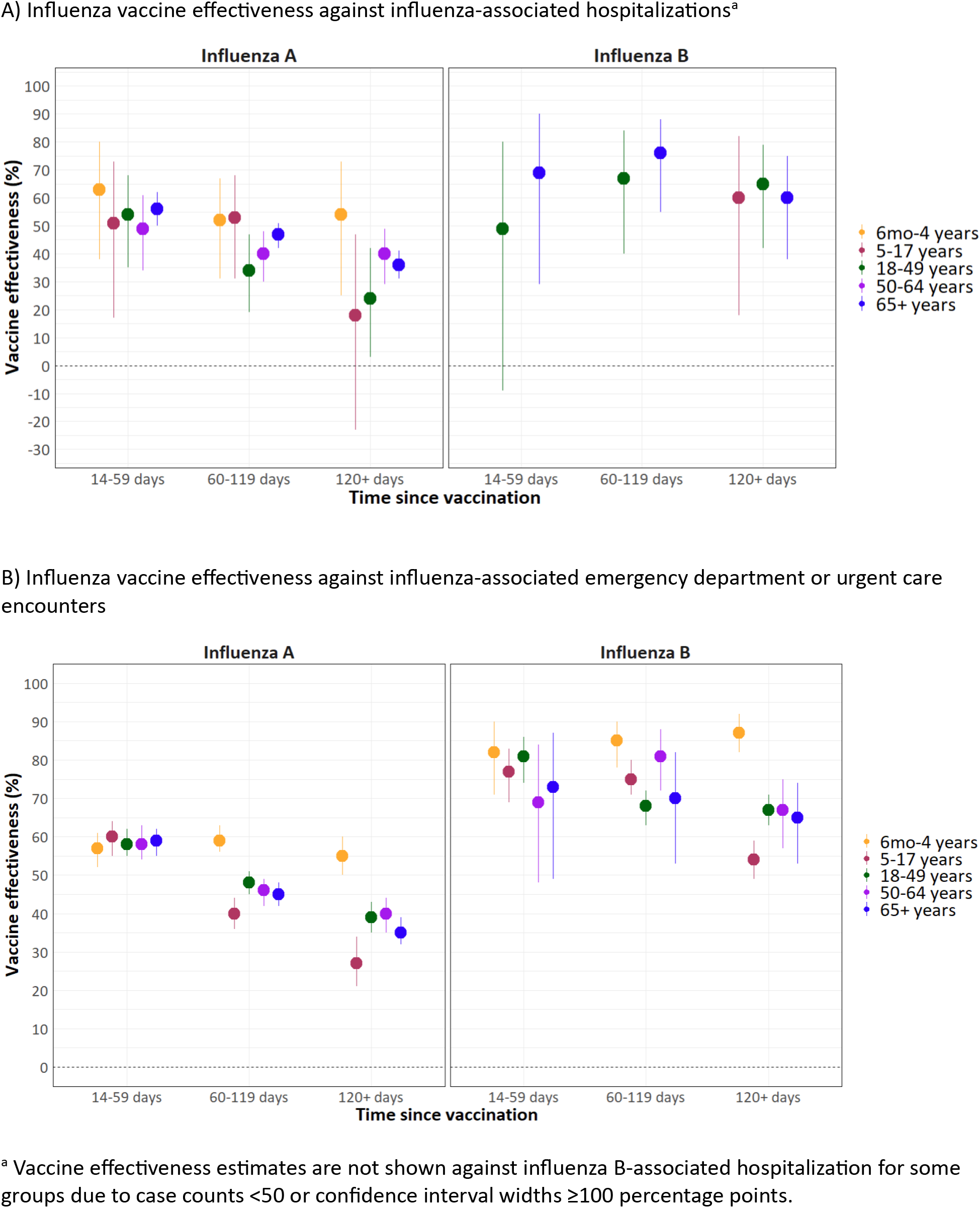
Influenza vaccine effectiveness against influenza-associated hospitalizations and emergency department or urgent care encounters by influenza type, age group, and time since vaccination — VISION, October 2024–April 2025

Among adults, VE against influenza-associated hospitalizations was 56% (95% CI: 51–60) at 14–59 days, 45% (95% CI: 42–49) at 60–119 days, and 37% (95% CI: 33–41) at ≥120 days after vaccination (Figure 2).

VE against ED/UC encounters was 60% (95% CI: 58–62) at 14–59 days, 48% (95% CI: 47–50) at 60–119 days, and 43% (95% CI: 41–45) at ≥120 days after vaccination. VE point estimates against influenza A-associated hospitalizations and ED/UC encounters were similar across adult age groups and generally declined with increasing time since vaccination (Figure 3, Supplemental Table 5), no consistent trends were observed against influenza B-associated hospitalizations and ED/UC encounters.

## DISCUSSION

Seasonal influenza vaccination provided protection against influenza-associated hospitalizations and ED/UC encounters among US children and adults during the 2024–25 season, with overall VE estimates ranging from 43–54%. Similar VE estimates were found among groups at higher risk of severe outcomes due to influenza, such as older adults, adults with immunocompromising conditions, and young children. Protection was also observed against critical illness, including ICU admission in children and ICU admission and death in adults. Although VE point estimates were highest during the first 2 months after vaccination, protection against hospitalization and ED/UC encounters was sustained for more than 4 months. Together, these findings support influenza vaccination as an important tool to reduce influenza-associated morbidity and mortality across the lifespan.

The 2024–25 influenza season was a high-severity season in all age groups, characterized by co-circulation of influenza A(H1N1)pdm09 and A(H3N2) viruses [1,3]. Co-circulation of both influenza A subtypes may have contributed to the elevated hospitalization rates observed, as well as the highest number of influenza-associated pediatric deaths observed since national reporting began in 2004 [2,14]. While both subtypes can cause severe illness, influenza A(H3N2)-predominant seasons have been associated with higher hospitalization rates among adults aged ≥65 years, and VE against A(H3N2) has historically been lower than that against A(H1N1) and B, particularly among older adults [15–17]. In the current analysis, influenza vaccination reduced the likelihood of influenza A-associated hospitalizations among children and adults, suggesting that vaccination provided protection against both A(H1N1)pdm09 and A(H3N2) influenza. This finding agrees with data from other US and Northern Hemisphere influenza VE studies [18–20]. Despite antigenic differences between circulating viruses and updated vaccine components, VE estimates from the current analysis against influenza A-associated hospitalizations and ED/UC encounters during the 2024–25 season (43–50%) were similar to those from the VISION Network during the 2023–24 season (37–49%) [11]. Using established networks that apply the same methodological approach to produce annual VE estimates has the benefit of allowing for comparisons between seasons.

In previous analyses, influenza vaccination has been associated with protection against a variety of critical outcomes, including severe and life-threatening pediatric influenza [6,21], pediatric influenza-associated deaths [22], and influenza-associated severe outcomes and deaths among adults [5,11,23]. Protection has also been found among adults with immunocompromising conditions at increased risk of influenza complications [24]. Results from the current analysis showing that influenza vaccination was associated with protection against influenza-associated ICU admission in children, ICU admission and death in adults, and hospitalizations in immunocompromised adults are consistent with these previous findings. Persistence of protection for >4 months after vaccination indicated that vaccine-elicited protection lasted through the 2024–25 influenza season. Lower VE estimates observed >4 months after vaccination were consistent with observations during the 2023-24 season [11].

Despite numerous analyses showing the benefits of influenza vaccination, vaccination coverage has decreased in most age groups since the start of the COVID-19 pandemic. From the 2019-20 season to the 2024-25 season, influenza vaccination coverage fell from 64% to 50% among US children aged 6 months–17 years and from 70% to 64% among US adults aged ≥65 years [25]. Decreasing influenza vaccination coverage has left an increasing number of US children and adults at higher risk of severe influenza-associated outcomes. However, despite continued declines in coverage, estimates indicate that influenza vaccination still prevented 10 million symptomatic illnesses, 5 million medical visits, 180,000 hospitalizations, and 12,000 deaths during the 2024–25 season [1]. Improving vaccination coverage could further reduce the burden of influenza-associated disease.

## Limitations

This analysis had several limitations. First, because VISION uses clinical testing to classify influenza case status and few cases are subtyped in routine clinical testing, we were unable to estimate VE by influenza A subtype. Second, although vaccination status was determined using multiple data sources, some influenza vaccine doses may not have been documented, resulting in misclassification of influenza vaccination status, however, misclassification would have occurred among both cases and controls, likely resulting in a bias toward the null and an underestimation of influenza VE. Third, due to limited data on influenza vaccination in previous seasons, we were unable to estimate VE among fully vaccinated versus partially vaccinated children aged 6 months–8 years [4]. Fourth, we did not account for influenza infection history in our VE models. Fifth, findings may not be representative of the entire US population.

## Conclusions

Influenza vaccination provided protection against influenza-associated hospitalizations and ED/UC encounters among US children and adults during the high-severity 2024–25 season. Increased uptake of influenza vaccine could reduce the burden of influenza and its complications.

## Data Availability

Data sharing agreements between the CDC and VISION partner institutions prohibit the CDC from making VISION data publicly available.

## Conflict of Interest Disclaimer

All authors have completed and submitted the International Committee of Medical Journal Editors form for disclosure of potential conflicts of interest. During the conduct of the study, all Kaiser Permanente Southern California Department of Research and Evaluation and Kaiser Permanente Northern California Division of Research-affiliated authors reported receiving contractual support from the Centers for Disease Control and Prevention (CDC) via payments made to their respective institutions. Additionally, all authors affiliated with Columbia University, HealthPartners Institute, Intermountain Health, Kaiser Permanente Center for Health Research, Regenstrief Institute, and University of Colorado Anschutz Medical Campus reported receiving contractual support from the CDC during the conduct of the study, via subcontracts from Westat, Inc. with payments made to their respective institutions. Unrelated to the submitted work, the following disclosures were reported from the past 36 months: SYT reports a contract from Pfizer. NPK reports support from Pfizer for a COVID-19 vaccine clinical trial and research support from Sanofi Pasteur, Merck, Pfizer, Moderna, Janssen, AstraZeneca, Seqirus, and GlaxoSmithKline. SJG reports contracts with National Institutes of Health (NIH) National Center for Advancing Translational Sciences and NIH National Institute of Mental Health. TCO received consulting fees from Regenstrief Institute and support for travel from Patient-Centered Outcomes Research Institute (PCORI) and Regenstrief Institute and has a current patent, PCT/US2018/047961. MBD reports grants from the Centers for Disease Control and Prevention (CDC) for the Vaccine Safety Datalink and Centers for Excellence in Newcomer Health. LSS reports contracts with GlaxoSmithKline, Moderna, Dynavax, and AstraZeneca. OZ reports grant number R01AI168373 with the National Institute of Allergy and Infectious Diseases and contracts with Moderna, Pfizer, and NIH. KBJ reports research support from Pfizer. BED reports a contract with Vaccine Safety Datalink Project, Contract No. 75D30122D15424. WFF reports a contract with Vaccine Safety Datalink Project, Contract No. 75D30122D15424. MAB was a speaker bureau participant – Innoviva Specialty Therapeutics - Oct 2023. AY reports a contract with University of Utah and Novavax. CEM reports support from NIH, Department of Defense, PCORI, AstraZeneca, and GlaxoSmithKline, was a Pri-Med lecturer, and is a member of the American Lung Association of Minnesota Board, Minnesota Department of Health Long COVID Advisory Committee, and Minnesota Department of Health Asthma Care Advisory Committee. TS is a member of CDC Advisory Committee on Immunization Practices Influenza Vaccine Work Group, chair of Utah Adult Immunization Coalition vaccine quality improvement and advocacy group, and a member of Utah Department of Health and Human Services Scientific Advisory Committee on Vaccines. KN reports support from NIH National Cancer Institute (NCI), National Heart, Lung, and Blood Institute (NHLBI), and Office of the Director. No other disclosures were reported.

## Disclaimer

The findings and conclusions of this report are those of the authors and do not necessarily reflect the official position of the Centers for Disease Control and Prevention.

## References

1. Centers for Disease Control and Prevention. 2024–2025 Influenza Season Summary: Severity, Disease Burden, and Burden Prevented. Available at: https://www.cdc.gov/flu-burden/php/data-vis-vac/2024-2025-prevented.html. Accessed 13 April 2026.

2. O’Halloran A, Habeck JW, Gilmer M, et al. Influenza-Associated Hospitalizations During a High Severity Season - Influenza Hospitalization Surveillance Network, United States, 2024-25 Influenza Season. MMWR Morb Mortal Wkly Rep 2025, 74:529–537.

3. Centers for Disease Control and Prevention. Influenza Activity in the United States during the 2024-25 Season and Composition of the 2025-26 Influenza Vaccine. 2025. Available at: https://www.cdc.gov/flu/whats-new/2025-2026-influenza-activity.html. Accessed 13 April 2026.

4. Grohskopf L, Ferdinands J, Blanton L, Broder K, Loehr J. Prevention and Control of Seasonal Influenza with Vaccines: Recommendations of the Advisory Committee on Immunization Practices — United States, 2024–25 Influenza Season. MMWR Recommendations and Reports 2024, 73:1–25.

5. Lewis NM, Zhu Y, Peltan ID, et al. Vaccine Effectiveness Against Influenza A–Associated Hospitalization, Organ Failure, and Death: United States, 2022–2023. Clinical Infectious Diseases 2024, 78:1056–1064.

6. Sumner KM, Sahni LC, Boom JA, et al. Estimated Vaccine Effectiveness for Pediatric Patients With Severe Influenza, 2015-2020. JAMA Network Open 2024, 7:e2452512–e2452512.

7. White EB, Grant L, Mak J, et al. Influenza Vaccine Effectiveness Against Illness and Asymptomatic Infection in 2022–2023: A Prospective Cohort Study. Clinical Infectious Diseases 2025, 80:893–900.

8. Chung JR, Price AM, Zimmerman RK, et al. Influenza Vaccine Effectiveness Against Medically Attended Outpatients Illness, United States, 2023–2024 Season. Clinical Infectious Diseases 2025, 81:e184–e191.

9. Centers for Disease Control and Prevention. CDC Seasonal Flu Vaccine Effectiveness Studies. Available at: https://www.cdc.gov/flu-vaccines-work/php/effectiveness-studies/index.html. Accessed 13 April 2026.

10. Centers for Disease Control and Prevention. VISION Vaccine Effectiveness Network. Available at: https://www.cdc.gov/flu-vaccines-work/php/vaccine-effectiveness/vision-network.html. Accessed 13 April 2026.

11. Tenforde MW, Reeves EL, Weber ZA, et al. Influenza Vaccine Effectiveness Against Hospitalizations and Emergency Department or Urgent Care Encounters for Children, Adolescents, and Adults During the 2023–2024 Season, United States. Clinical Infectious Diseases 2025, 81:667–678.

12. Doll MK, Pettigrew SM, Ma J, Verma A. Effects of Confounding Bias in Coronavirus Disease 2019 (COVID-19) and Influenza Vaccine Effectiveness Test-Negative Designs Due to Correlated Influenza and COVID-19 Vaccination Behaviors. Clinical Infectious Diseases 2022, 75:e564–e571.

13. Lewis NM, Harker EJ, Leis A, et al. Assessment and mitigation of bias in influenza and COVID-19 vaccine effectiveness analyses — IVY Network, September 1, 2022–March 30, 2023. Vaccine 2025, 43:126492.

14. Centers for Disease Control and Prevention. FluView Interactive: Influenza-Associated Pediatric Mortality. Available at: https://gis.cdc.gov/GRASP/Fluview/PedFluDeath.html. Accessed 13 April 2026.

15. Budd AP, Beacham L, Smith CB, et al. Birth Cohort Effects in Influenza Surveillance Data: Evidence That First Influenza Infection Affects Later Influenza-Associated Illness. The Journal of Infectious Diseases 2019, 220:820–829.

16. Belongia EA, Simpson MD, King JP, et al. Variable influenza vaccine effectiveness by subtype: a systematic review and meta-analysis of test-negative design studies. The Lancet Infectious Diseases 2016, 16:942–951.

17. Okoli GN, Racovitan F, Abdulwahid T, Righolt CH, Mahmud SM. Variable seasonal influenza vaccine effectiveness across geographical regions, age groups and levels of vaccine antigenic similarity with circulating virus strains: A systematic review and meta-analysis of the evidence from test-negative design studies after the 2009/10 influenza pandemic. Vaccine 2021, 39:1225–1240.

18. Frutos AM. Interim estimates of 2024–2025 seasonal influenza vaccine effectiveness—four vaccine effectiveness networks, United States, October 2024–February 2025. MMWR Morbidity and mortality weekly report 2025, 74.

19. Rose AM, Lucaccioni H, Marsh K, et al. Interim 2024/25 influenza vaccine effectiveness: eight European studies, September 2024 to January 2025. Eurosurveillance. 2025, 30:2500102.

20. Separovic L, Zhan Y, Kaweski SE, et al. Interim estimates of vaccine effectiveness against influenza A(H1N1)pdm09 and A(H3N2) during a delayed influenza season, Canada, 2024/25. Eurosurveillance. 2025, 30:2500059.

21. Olson SM, Newhams MM, Halasa NB, et al. Vaccine Effectiveness Against Life-Threatening Influenza Illness in US Children. Clinical Infectious Diseases 2022, 75:230–238.

22. Flannery B, Reynolds SB, Blanton L, et al. Influenza Vaccine Effectiveness Against Pediatric Deaths: 2010–2014. Pediatrics 2017, 139:e20164244.

23. Lewis NM, Harker EJ, Cleary S, et al. Vaccine Effectiveness Against Influenza A(H1N1), A(H3N2), and B-Associated Hospitalizations, United States, 1 September 2023 to 31 May 2024. The Journal of Infectious Diseases 2025, 232:e626–e636.

24. Scott J, Abers MS, Marwah HK, et al. Updated Evidence for Covid-19, RSV, and Influenza Vaccines for 2025-2026. N Engl J Med 2025, 393:2221–2242.

25. Centers for Disease Control and Prevention. Influenza Vaccination Coverage for Persons 6 Months and Older. Available at: https://www.cdc.gov/fluvaxview/interactive/general-population-coverage.html. Accessed 13 April 2026.

