## Supplemental Materials for "Influenza vaccine effectiveness against influenza-associated hospitalizations and emergency department or urgent care encounters among children and adults — United States, 2024–25 season"

**Supplemental Table 1.** ICD-10 Codes Used to Define Acute Respiratory Illness

| **Acute Respiratory Illness Description** | **ICD-10 Codes** |
| --- | --- |
| Influenza Pneumonia | J09.X1, J10.0*, J11.0* |
| Other Viral Pneumonia | J12.* (excluding J12.81 and J12.82) |
| Bacterial Pneumonia | J13, J14, J15.*, J16.*, J17, J18.* |
| Influenza Disease | J09.*, J10.1, J10.2, J10.8*, J11.1, J11.2, J11.8* (excluding J09.X1) |
| Acute Respiratory Distress Syndrome | J80 |
| COPD Exacerbation | J44.1[a] |
| Asthma Exacerbation | J45.21, J45.22, J45.31, J45.32, J45.41, J45.42, J45.51, J45.52, J45.901, J45.902 |
| Respiratory Failure | J96.0*, J96.2*, R09.2, J96.9* |
| Other Acute Lower Respiratory Tract Infections | B97.4, J20.*, J21.*, J22, J40, J44.0[a], J41.*, J42, J43.*, J47.*, J85.*, J86.* |
| Sinusitis | J01.*, J32.*[a] |
| Acute Upper Respiratory Tract Infections | J00.*, J02.*, J03.*, J04.*, J05.*, J06.* |
| Acute Respiratory Illness Signs and Symptoms | R04.2, R05, R05.1, R05.2, R05.4, R05.8, R05.9, R06.00, R06.02, R06.03, R06.1, R06.2, R06.8, R06.81, R06.82, R06.89, R07.1, R09.0*, R09.1, R09.2, R09.3, R09.8* |

Note: All ICD-10 codes with * include all child codes under the specific parent code.

[a]Denotes ICD codes used only for adults aged ≥18 years.

**Supplemental Table 2.** Dates of inclusion in influenza vaccine effectiveness analyses by VISION site and care setting[a]

| Site | Hospitalizations | | Emergency Department or Urgent Care Encounters | |
|---|---|---|---|---|
| | Start Date | End Date | Start Date | End Date |
| Site A | 10/13/2024 | 4/29/2025 | 10/5/2024 | 4/30/2025 |
| Site B | 11/8/2024 | 4/28/2025 | 10/3/2024 | 4/30/2025 |
| Site C | 10/8/2024 | 4/30/2025 | 10/1/2024 | 4/30/2025 |
| Site D | 10/3/2024 | 4/29/2025 | 10/1/2024 | 4/30/2025 |
| Site E | 10/19/2024 | 4/29/2025 | 10/3/2024 | 4/30/2025 |
| Site F | 10/1/2024 | 4/30/2025 | 10/2/2024 | 4/30/2025 |
| Site G | 10/2/2024 | 4/30/2025 | 10/1/2024 | 4/30/2025 |

Abbreviations: VISION = Virtual SARS-CoV-2, Influenza, and Other respiratory viruses Network

[a] Influenza vaccine effectiveness analyses included acute respiratory illness-associated encounters from the date of the first influenza-positive case on or after October 1, 2024 by VISION site and setting to the date of the last influenza-positive case on or before April 30, 2025 by VISION site and setting.

**Supplemental Table 3.** ICD-10 Codes Used to Define Underlying Medical Conditions and Immunocompromising Conditions

| **Underlying Medical Condition** | **ICD-10 Codes** |
|---|---|
| Respiratory Condition | J45.*, J40.[a], J41.*[a], J42.[a], J43.*[a], J44.*[a], D86.0, E88.01, J47.*, J60[a], J61[a], J62.*[a], J63.*[a], J64[a], J65[a], J66.*, J67.0, J67.1, J67.2, J67.3, J67.4, J67.5, J67.6, J67.7, J67.8, J68.*, J70.*, J81.1, J84.*, J95.*, J96.1*, J99., P26.*, P27.*, B39.*, B40.1, B40.2, B41.0, B44.0, B44.1 , B45.*, B46.0, A15.*, A31.0, G47.3*[b], P27.0[b], P27.1[b], P27.8[b], P27.9[b], E84.*, Q33.*[b],Q34.*[b], Q30.*[b], Q31.*[b], Q32.*[b], Q39.*[b], Q79.0[b], J45.901[b], J45.902[b], J45.909[b] |
| Cardiovascular Condition | I50.*, I21.*, I22.*, I23.*, I24.*, I25.*, I10., I11.*, I13.*, I15.*, I01.*, I02.0, I09.*, I27.*, I28.*, I31.*, I42.*, I43., I44.*, I46.*, I51.0, I51.1, I51.2, I51.3, I51.5, I51.7, I51.8*, I51.9, I52., I97.0, I97.1*, M31.0, M31.1*, M31.2, M31.4, M31.6, M31.7, M31.8, M31.9, Z95.*, Z98.61, I71.*, I72.*, I73.*, I74.*, I75.*, I79.*, I26.*, I05.*, I06.*, I07.*, I08.*, I34.*, I35.*, I36.*, I37.*, I48.*, I50.9, I42.9, Q20.*[a], Q21.*, Q22.*, Q23.*, Q24.*, Q25.*, Q26.*, Q27.0, Q27.3*, Q27.4, Q27.8, Q27.9, Q28.*, P29.30, Q89.3[a] |
| Cerebrovascular Condition | I60.*, I61.*, I63.*, I62.*, I68.*, I69.* |
| Neurologic and/or Musculoskeletal Condition | F01.*[a], F02.*[a], F03.*[a], G30.*[a], H49.81*, M12.0*, M36.0, E75.02, E75.19, E75.4, F71., F72., F73., F84.2, G10., G11.*, G12.*, G13.*, G14., G20.*, G21.*, G23.*, G24.*, G25.*, G26., G31.*, G32.*, G35., G36.*, G37.*, G40.*, G45.*, G46.*, G60.*, G61.*, G62.*, G63., G64., G70.*, G71.*, G73.*, G80.*, G81.*, G82.*, G83.*, G90.3, G91.*, G93.*, G94., G95.*, G99.2, P91.*, Q00.*, Q01.*, Q02., Q03.*, Q04.*, Q05.*, Q06.*, Q07.*, Q76.*, Q77.*, Q78.*, Q79.1, Q79.2, Q79.3, Q79.4, Q79.5*, Q79.6*, Q79.8, Q79.9, Q85.*, Q87.4*, Q91.*, Q92.*, Q93.*, Q96.*, R41.*, R53.2, R54., Q90.*, R56.*[b], P90[b], P91.0[b], P52.*[b], E70.*[b], E71.*[b], E72.*[b], E74.*[b], E75.2*[b], E76.*[b], E77.*[b], E78.*[b], E79.*[b], E80.*[b], P70.*[b], P71.*[b], P72.*[b], P74.*[b], P94.*[b] |
| Hematologic Condition | D55.*, D56.0, D56.1, D56.2, D56.4, D56.5, D56.8, D56.9, D57.0*, D57.1, D57.2*, D57.4*, D57.8*, D58.*, D59.*, D60.*, D61.*, D64.0, D64.1, D64.2, D64.3, D64.4, D64.8*, D65, D66, D67, D68.* |
| Endocrine Condition | E10.*, E11.*, E08.*, E09.*, E13.*, E00.*, E01.*, E03.*, E05.*, E06.*, E15. , E16.*, E20.*, E21.*, E22.*, E23.*, E24.*, E25.*, E26.*, E27.*, E28.*, E29.*, E31.*, E32.*, E34.*, E70.*[a], E71.*[a], E72.*[a], E74.*[a], E75.2*[a], E76.*[a], E77.*[a], E78.*[a], E79.*[a], E80.*[a], E83.*, E85.*, E88.02, E88.09, E88.1, E88.2, E88.3, E88.4*, E88.8*, E88.9 |
| Renal Disease | I12.*, I13.* N01.*, N02.*, N03.*, N04.*, N05.*, N06.*, N07.*, N08., N11.*, N14.*, N15.*, N16., N18.*, N25.*, N26.*, N28.*, Q27.1, Q27.2, Q60.*, Z49.*, Z91.15*, Z94.0, Z99.2 |
| Gastrointestinal Condition | B18.*, I81, I85.*, K70.*, K71.*, K72.*, K73.*, K74.*, K75.*, K76.*, K77., K50.*, K51.*, K52.* |
| Other Underlying Conditions | P07.*[b], R62.*[b], Z93.0[b], Z93.1[b], Z99.0[b], Z99.11[b], Z99.81[b], Z99.89[b] |

| | |
|---|---|
| Immunocompromising Conditions | **Hematologic Malignancy**: C81.*, C82.*, C83.*, C84.*, C85.*, C86.*, C88.*, C90.*, C91.*, C92.*, C93.*, C94.*, C95.*, C96.*, D46.*, D61.0*, D70.0, D61.2, D61.9, D71.*<br>**Solid Malignancy**: C00.*, C01., C02.*, C03.*, C04.*, C05.*, C06.*, C07., C08.*, C09.*, C10.*, C11.*, C12., C13.*, C14.*, C15.*, C16.*, C17.*, C18.*, C19., C20., C21.*, C22.*, C23., C24.*, C25.*, C26.*, C30.*, C31.*, C32.*, C33, C34.*, C37, C38.*, C39.*, C40.*, C41.*, C43.*, C45.*, C46.*, C47.*, C48.*, C49.*, C50.*, C51.*, C52, C53.*, C54.*, C55, C56.*, C57.*, C58, C60.*, C61, C62.*, C63.*, C64.*, C65.*, C66.*, C67.*, C68.*, C69.*, C70.*, C71.*, C72.*, C73, C74.*, C75.*, C76.*, C77.*, C78.*, C79.*, C7A.*, C7B.*, C80.*, Z51.0, Z51.1*, C4A.*<br>**Solid Organ or Bone Marrow Transplant**: T86.0*, T86.1*, T86.2*, T86.3*, T86.4*, T86.5, T86.81*, T86.85*, D47.Z1, Z48.2*, Z94.*, Z98.85<br>**Rheumatologic/inflammatory Disorders:** D86.*, E85.1, E85.2, E85.3, E85.4, E85.8*, E85.9, G35.*, J67.9, L40.54, L40.59, L93.0, L93.2, L94.*, M05.*, M06.*, M07.*, M08.*, M30.*, M31.3*, M31.5, M32.*, M33.*, M34.*, M35.3, M35.89, M35.9, M46.0*, M46.1, M46.8*, M46.9*<br>**Other Intrinsic Immune Condition or Immunodeficiency**: D27.9, D72.89, D80.*, D81.0, D81.1, D81.2, D81.4, D81.5, D81.6, D81.7, D81.8*, D81.9, D82.*, D83.*, D84.*, D89.0, D89.1, D89.3, D89.4*, D89.8*, D89.9, K70.3*, K70.4*, K72.*, K74.3, K74.4, K74.5, K74.6*, N04.*, R18.0<br>**HIV/AIDS:** B20., B97.35, O98.7*, Z21, E88.14 |

Abbreviations: ICD = International Classification of Diseases

Note: All ICD-10 codes with * include all child codes under the specific parent code.

[a]Denotes ICD codes used only for adults aged ≥18 years.

[b]Denotes ICD codes used only for children aged <18 years.

**Supplemental Figure 1.** Exclusion flow diagrams for hospitalizations and emergency department or urgent care encounters

A) Exclusion flow diagram for hospitalizations

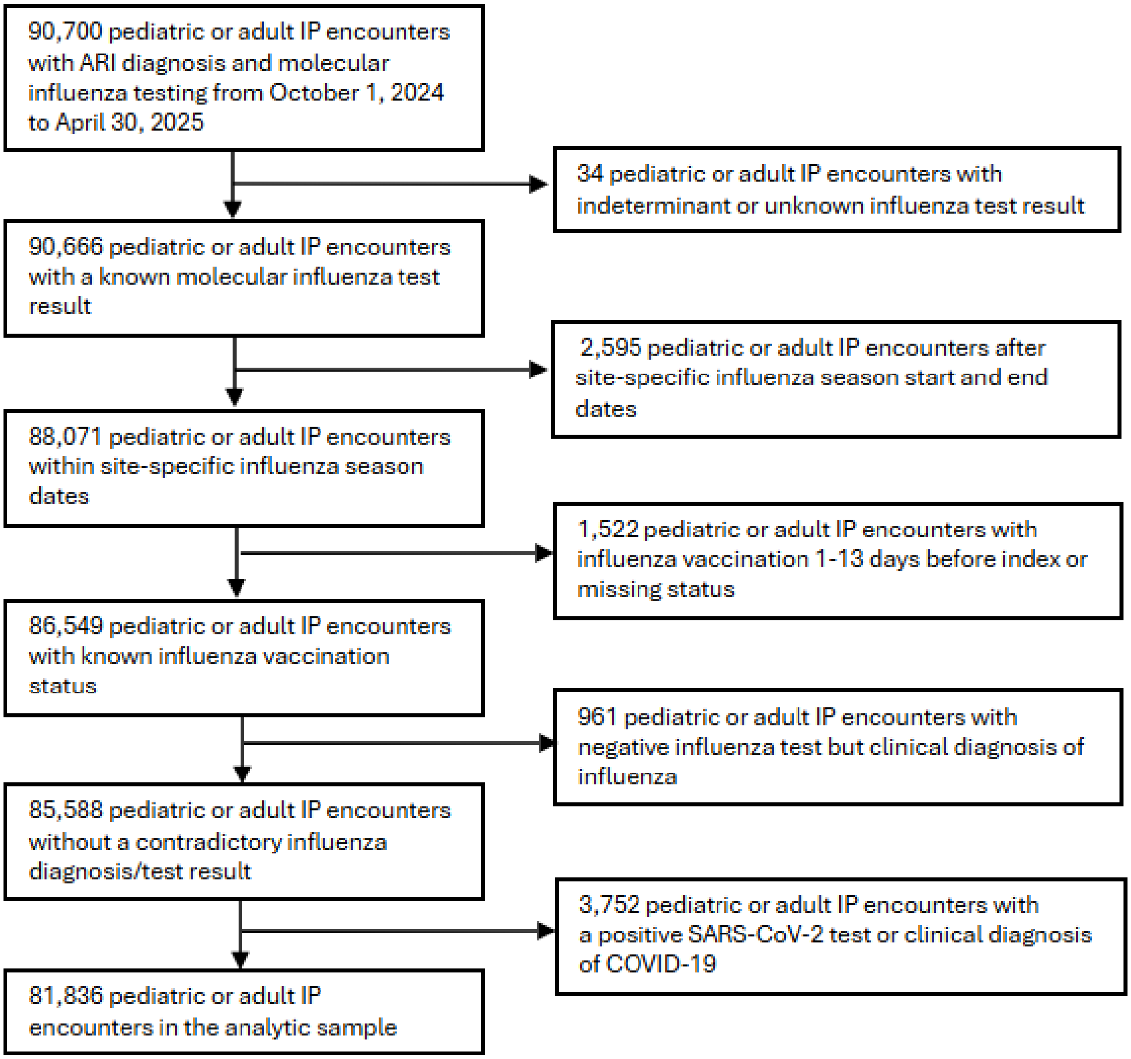

B) Exclusion flow diagram for emergency department or urgent care encounters

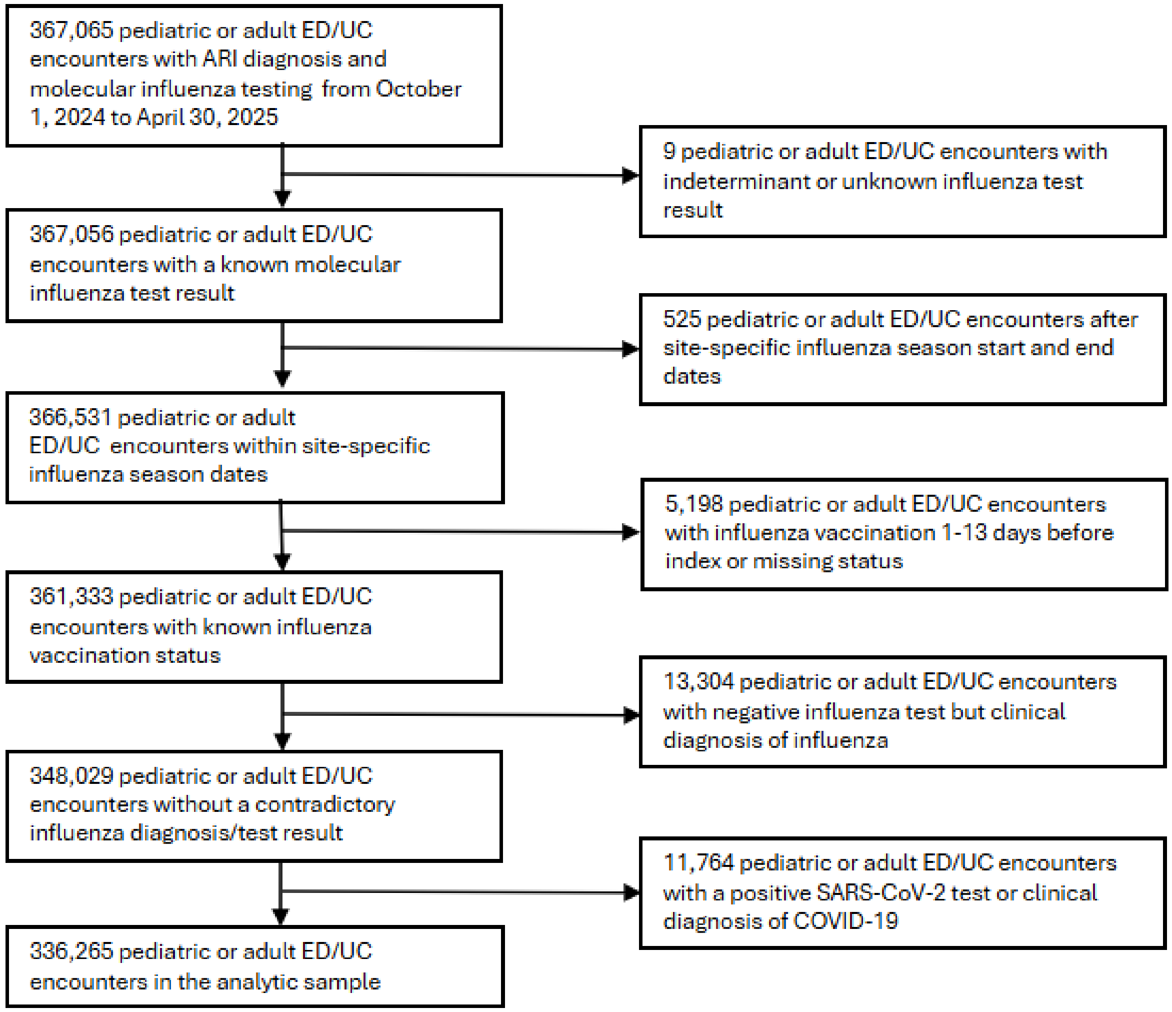


Abbreviations: ARI = acute respiratory illness; ED/UC = emergency department or urgent care; IP = inpatient

**Supplemental Figure 2**. Influenza-associated medical encounters and percent positivity by week, age group, and setting — VISION, October 2024–April 2025

A) Influenza-positive hospitalizations and percent positivity by week among children aged 6 months–17 years

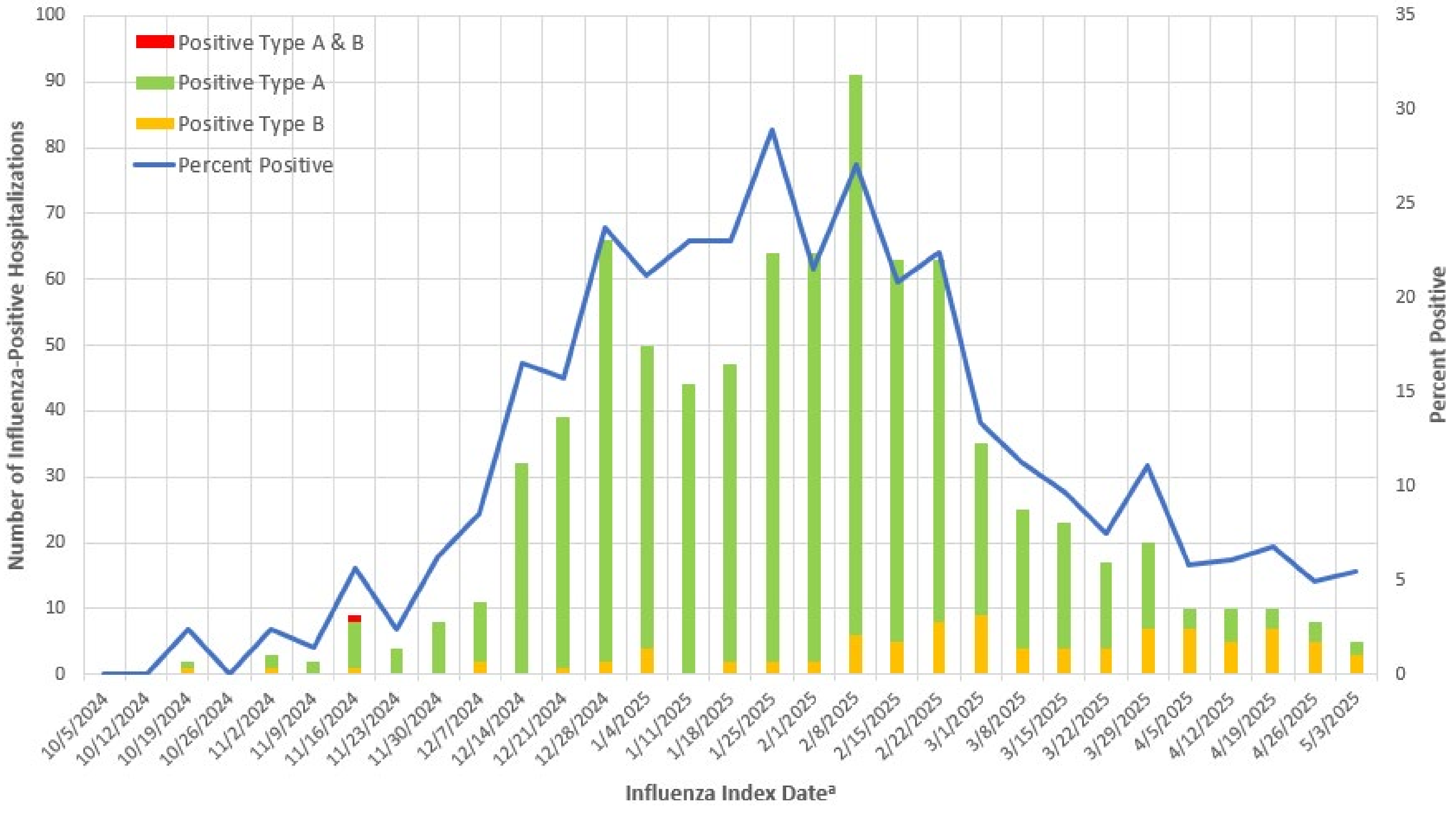

B) Influenza-positive emergency department or urgent care encounters and percent positivity by week among children aged 6 months–17 years

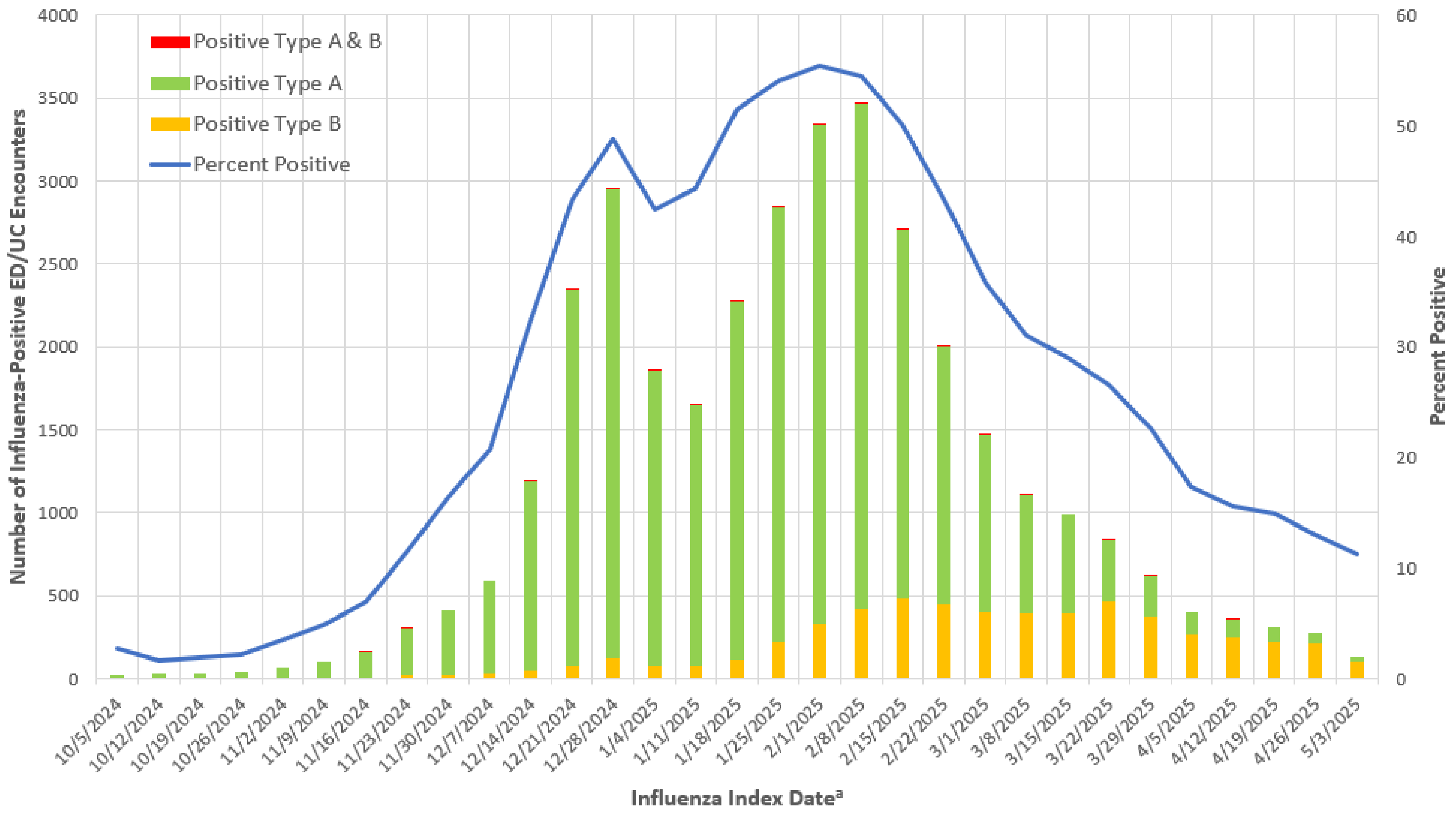

C) Influenza-positive hospitalizations and percent positivity by week among adults aged ≥18 years

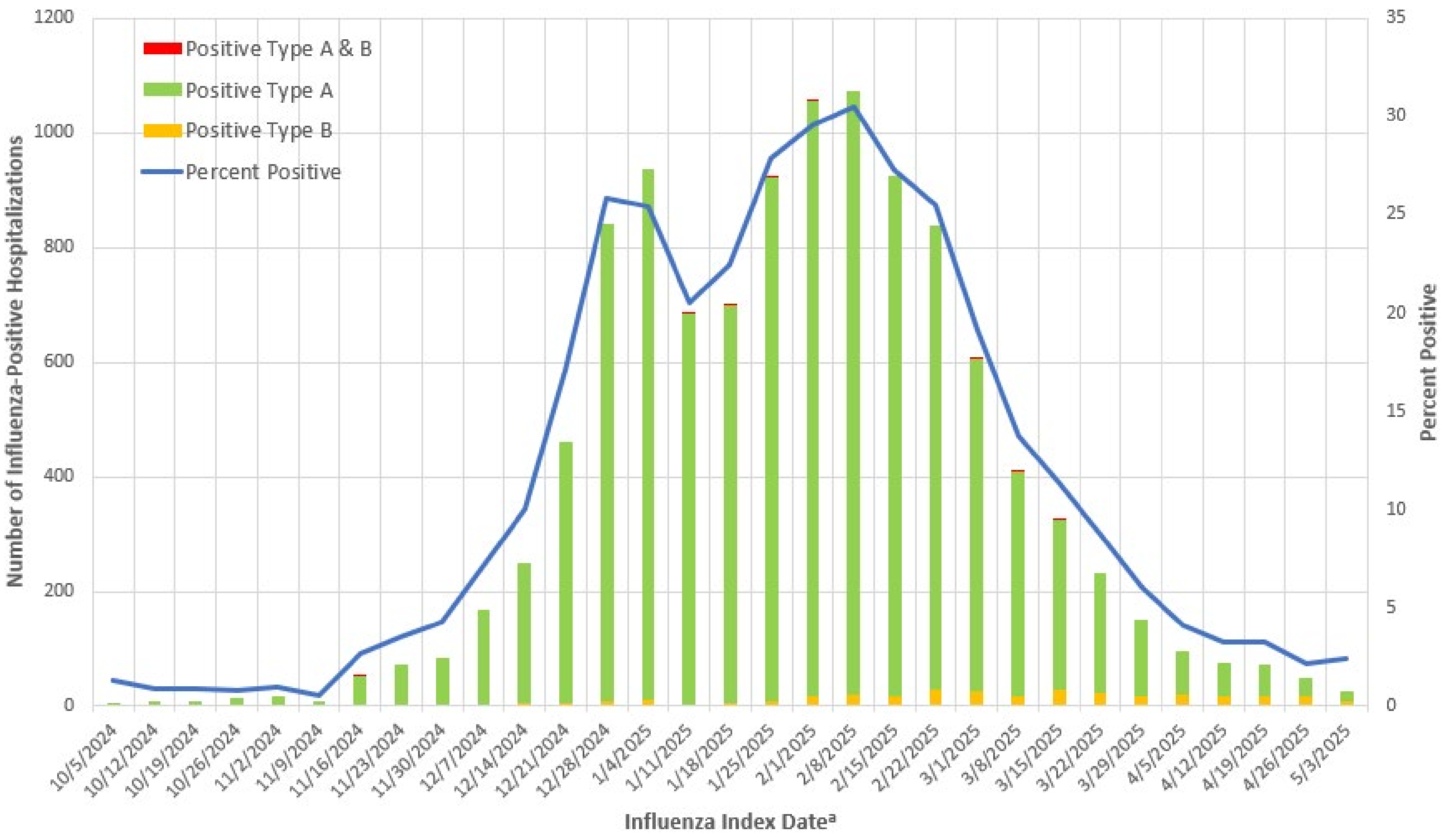

D) Influenza-positive emergency department or urgent care encounters and percent positivity by week among adults aged ≥18 years

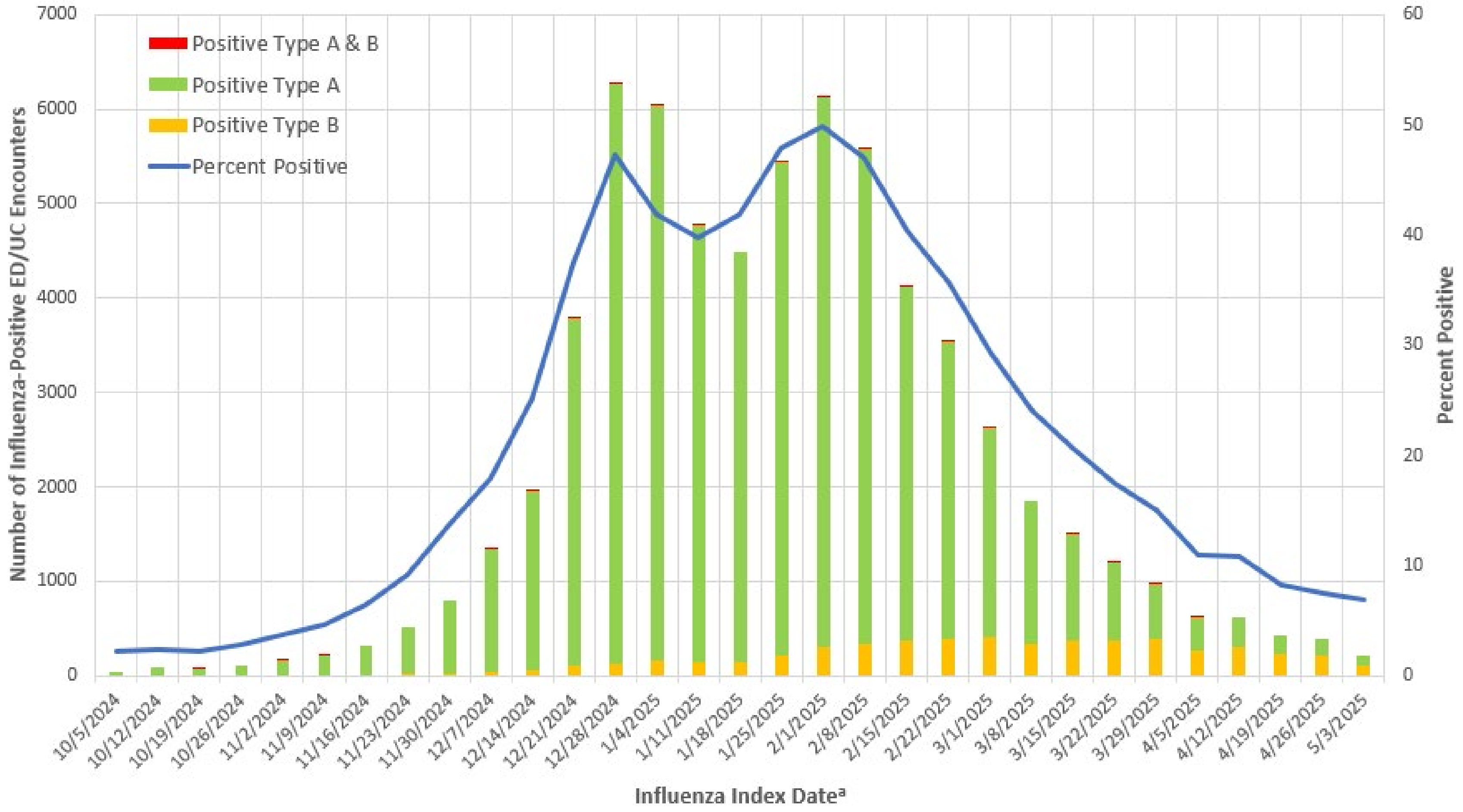


Abbreviations: ED/UC = emergency department or urgent care

[a] Influenza index date was defined as the earlier date of either the most recent influenza test or the encounter date.

**Supplemental Table 4**. Influenza vaccine product type received by site and age group — VISION, October 2024–April 2025[a]

| | N (row %)[b] | | | | | | | N (row %)[c] |
|---|---|---|---|---|---|---|---|---|
| Age Group and Site | Live Attenuated (FluMist) | Standard-Dose Inactivated (Afluria, Fluarix, FluLaval, Fluzone) | Cell Culture-Based (Flucelvax) | Recombinant (Flublok) | High-Dose Inactivated (Fluzone High-Dose) | Adjuvanted (Fluad) | Other | Unknown/ Missing product type |
| Total | 1,100 (0.8) | 64,215 (44.3) | 7,011 (4.8) | 2,531 (1.7) | 51,160 (35.3) | 16,701 (11.5) | 2,257 (1.6) | 6,963 (4.6) |
| 6 months–17 years | 1,050 (4.0) | 22,384 (84.5) | 2,599 (9.8) | 2 (0.0) | 5 (0.0) | 5 (0.0) | 435 (1.6) | 1,066 (3.9) |
| Site A | 0 (0.0) | 687 (99.6) | 1 (0.1) | 0 (0.0) | 0 (0.0) | 0 (0.0) | 2 (0.3) | 3 (0.4) |
| Site B | 161 (10.7) | 369 (24.6) | 969 (64.5) | 0 (0.0) | 0 (0.0) | 0 (0.0) | 3 (0.2) | 2 (0.1) |
| Site C | 189 (5.1) | 2,803 (75.5) | 433 (11.7) | 0 (0.0) | 1 (0.0) | 1 (0.0) | 285 (7.7) | 930 (20.0) |
| Site D | 72 (2.0) | 3,016 (82.8) | 469 (12.9) | 2 (0.1) | 0 (0.0) | 1 (0.0) | 83 (2.3) | 41 (1.1) |
| Site E | 24 (1.0) | 1,989 (84.4) | 317 (13.4) | 0 (0.0) | 1 (0.0) | 1 (0.0) | 26 (1.1) | 3 (0.1) |
| Site F | 13 (0.2) | 5,898 (93.4) | 392 (6.2) | 0 (0.0) | 0 (0.0) | 1 (0.0) | 10 (0.2) | 13 (0.2) |
| Site G | 591 (7.2) | 7,622 (92.3) | 18 (0.2) | 0 (0.0) | 3 (0.0) | 1 (0.0) | 26 (0.3) | 74 (0.9) |
| 18–64 years | 43 (0.1) | 37,534 (83.7) | 4,152 (9.3) | 1,730 (3.9) | 480 (1.1) | 115 (0.3) | 812 (1.8) | 2,831 (5.9) |
| Site A | 0 (0.0) | 1,684 (94.6) | 44 (2.5) | 30 (1.7) | 12 (0.7) | 0 (0.0) | 11 (0.6) | 21 (1.2) |
| Site B | 18 (0.6) | 1,051 (35.9) | 1,664 (56.8) | 100 (3.4) | 8 (0.3) | 14 (0.5) | 76 (2.6) | 20 (0.7) |
| Site C | 8 (0.3) | 2,233 (72.2) | 303 (9.8) | 299 (9.7) | 53 (1.7) | 49 (1.6) | 149 (4.8) | 1,408 (31.3) |
| Site D | 2 (0.0) | 4,593 (69.8) | 842 (12.8) | 755 (11.5) | 38 (0.6) | 6 (0.1) | 342 (5.2) | 254 (3.7) |
| Site E | 0 (0.0) | 2,534 (60.9) | 998 (24.0) | 505 (12.1) | 53 (1.3) | 12 (0.3) | 58 (1.4) | 53 (1.3) |
| Site F | 0 (0.0) | 9,718 (97.2) | 126 (1.3) | 10 (0.1) | 61 (0.6) | 29 (0.3) | 51 (0.5) | 179 (1.8) |
| Site G | 15 (0.1) | 15,721 (96.3) | 175 (1.1) | 31 (0.2) | 255 (1.6) | 5 (0.0) | 125 (0.8) | 896 (5.2) |
| ≥65 years | 7 (0.0) | 4,297 (5.8) | 260 (0.4) | 799 (1.1) | 50,675 (68.8) | 16,581 (22.5) | 1,010 (1.4) | 3,066 (4.0) |
| Site A | 0 (0.0) | 90 (2.6) | 26 (0.7) | 31 (0.9) | 3,321 (94.7) | 34 (1.0) | 6 (0.2) | 32 (0.9) |
| Site B | 0 (0.0) | 62 (1.2) | 70 (1.4) | 21 (0.4) | 1,659 (32.9) | 3,115 (61.8) | 117 (2.3) | 53 (1.0) |

| | | | | | | | | |
|---|---|---|---|---|---|---|---|---|
| Site C | 0 (0.0) | 113 (2.7) | 17 (0.4) | 56 (1.3) | 3,106 (74.8) | 735 (17.7) | 125 (3.0) | 1,680 (28.8) |
| Site D | 4 (0.1) | 143 (2.9) | 19 (0.4) | 99 (2.0) | 3,860 (77.1) | 551 (11.0) | 329 (6.6) | 170 (3.3) |
| Site E | 0 (0.0) | 544 (6.1) | 83 (0.9) | 459 (5.2) | 6,055 (68.0) | 1,570 (17.6) | 193 (2.2) | 103 (1.1) |
| Site F | 0 (0.0) | 1,990 (7.8) | 15 (0.1) | 60 (0.2) | 12,933 (50.5) | 10,454 (40.8) | 150 (0.6) | 402 (1.5) |
| Site G | 3 (0.0) | 1,355 (6.3) | 30 (0.1) | 73 (0.3) | 19,741 (92.2) | 122 (0.6) | 90 (0.4) | 626 (2.8) |

[a] Influenza vaccine product types are defined as follows: Live Attenuated (CVX 111), Standard-Dose Inactivated (CVX 140,141), Cell Culture-Based (CVX 153, 320), Recombinant (CVX 155), High-Dose Inactivated (CVX 135), Adjuvanted (CVX 168), Other (CVX 15, 16, 123, 125, 126, 127, 128, 144, 149, 150, 158, 160, 161, 166, 171, 185, 186, 194, 197, 200, 201, 202, 205, 231, 321, 322, 323), Unknown/Missing product type (CVX 88, 151, or missing/unknown).

[b] The denominators for these row percentages only include influenza vaccinations with a known vaccine product type.

[c] The denominators for these row percentages include all influenza vaccinations with known and unknown vaccine product types

**Supplemental Table 5.** Influenza vaccine effectiveness against influenza-associated hospitalizations and emergency department or urgent care encounters by influenza type, age group, and time since vaccination — VISION, October 2024–April 2025

| | Hospitalizations[a] | | | Emergency Department or Urgent Care Encounters | | |
|---|---|---|---|---|---|---|
| | Cases<br><br>Number vaccinated / total (%) | Controls<br><br>Number vaccinated / total (%) | Vaccine effectiveness[b] | Cases<br><br>Number vaccinated / total (%) | Controls<br><br>Number vaccinated / total (%) | Vaccine effectiveness[b] |
| **Influenza A** | | | | | | |
| By age and time since vaccination | | | | | | |
| **6 months–4 yrs** | | | | | | |
| 14-59 days | 15/246 (6%) | 324/2017 (16%) | 63 (38-80) | 483/8761 (6%) | 3371/25992 (13%) | 57 (52-61) |
| 60-119 days | 38/269 (14%) | 415/2108 (20%) | 52 (31-67) | 904/9182 (10%) | 4618/27239 (17%) | 59 (56-63) |
| ≥120 days | 20/251 (8%) | 405/2098 (19%) | 54 (25-73) | 489/8767 (6%) | 3813/26434 (14%) | 55 (50-60) |
| **5-17 yrs** | | | | | | |
| 14-59 days | 17/351 (5%) | 161/1614 (10%) | 51 (17-73) | 547/16383 (3%) | 2194/27831 (8%) | 60 (55-64) |
| 60-119 days | 41/375 (11%) | 223/1676 (13%) | 53 (31-68) | 1724/17560 (10%) | 2789/28426 (10%) | 40 (36-44) |
| ≥120 days | 36/370 (10%) | 265/1718 (15%) | 18 (-23-47) | 1001/16837 (6%) | 2871/28508 (10%) | 27 (21-34) |
| **18-49 yrs** | | | | | | |
| 14-59 days | 47/1214 (4%) | 529/6350 (8%) | 54 (35-68) | 795/28493 (3%) | 4514/60723 (7%) | 58 (55-62) |
| 60-119 days | 183/1350 (14%) | 761/6582 (12%) | 34 (19-47) | 2639/30337 (9%) | 6411/62620 (10%) | 48 (45-51) |
| ≥120 days | 117/1284 (9%) | 851/6672 (13%) | 24 (3-42) | 1628/29326 (6%) | 6636/62845 (11%) | 39 (35-43) |
| **50-64 yrs** | | | | | | |

| | | | | | | |
|---|---|---|---|---|---|---|
| 14-59 days | 79/1667 (5%) | 1030/8653 (12%) | 49 (34-61) | 479/9880 (5%) | 2981/22544 (13%) | 58 (54-63) |
| 60-119 days | 310/1898 (16%) | 1597/9220 (17%) | 40 (30-48) | 1878/11279 (17%) | 4655/24218 (19%) | 46 (42-49) |
| ≥120 days | 233/1821 (13%) | 1863/9486 (20%) | 40 (29-49) | 1424/10825 (13%) | 5298/24861 (21%) | 40 (35-44) |
| **≥65 yrs** | | | | | | |
| 14-59 days | 330/4067 (8%) | 5670/24055 (24%) | 56 (50-62) | 711/7707 (9%) | 8175/30020 (27%) | 59 (55-62) |
| 60-119 days | 1515/5252 (29%) | 8995/27380 (33%) | 47 (42-51) | 3705/10701 (35%) | 13727/35572 (39%) | 45 (42-48) |
| ≥120 days | 1526/5263 (29%) | 11769/30154 (39%) | 36 (31-41) | 3309/10305 (32%) | 17098/38943 (44%) | 35 (32-39) |
| **Influenza B** | | | | | | |
| By age and time since vaccination | | | | | | |
| **6 months – 4 yrs** | | | | | | |
| 14-59 days | 1/23 (4%) | 324/2017 (16%) | -- | 15/733 (2%) | 3371/25992 (13%) | 82 (71-90) |
| 60-119 days | 1/23 (4%) | 415/2108 (20%) | -- | 26/744 (3%) | 4618/27239 (17%) | 85 (78-90) |
| ≥120 days | 1/23 (4%) | 405/2098 (19%) | -- | 30/748 (4%) | 3813/26434 (14%) | 87 (82-92) |
| **5-17 yrs** | | | | | | |
| 14-59 days | 1/53 (2%) | 161/1614 (10%) | -- | 60/4193 (1%) | 2194/27831 (8%) | 77 (69-83) |
| 60-119 days | 5/57 (9%) | 223/1676 (13%) | -- | 154/4287 (4%) | 2789/28426 (10%) | 75 (71-80) |
| ≥120 days | 9/61 (15%) | 265/1718 (15%) | 60 (18-82) | 474/4607 (10%) | 2871/28508 (10%) | 54 (49-59) |
| **18-49 yrs** | | | | | | |
| 14-59 days | 7/155 (5%) | 529/6350 (8%) | 49 (-9-80) | 41/4254 (1%) | 4514/60723 (7%) | 81 (74-86) |
| 60-119 days | 11/159 (7%) | 761/6582 (12%) | 67 (40-84) | 186/4399 (4%) | 6411/62620 (10%) | 68 (63-72) |

| | | | | | | |
|---|---|---|---|---|---|---|
| ≥120 days | 21/169 (12%) | 851/6672 (13%) | 65 (42-79) | 365/4578 (8%) | 6636/62845 (11%) | 67 (63-71) |
| **50-64 yrs** | | | | | | |
| 14-59 days | 2/40 (5%) | 1030/8653 (12%) | -- | 12/424 (3%) | 2981/22544 (13%) | 69 (48-84) |
| 60-119 days | 1/39 (3%) | 1597/9220 (17%) | -- | 21/433 (5%) | 4655/24218 (19%) | 81 (72-88) |
| ≥120 days | 10/48 (21%) | 1863/9486 (20%) | -- | 75/487 (15%) | 5298/24861 (21%) | 67 (57-75) |
| **≥65 yrs** | | | | | | |
| 14-59 days | 3/58 (5%) | 5670/24055 (24%) | 69 (29-90) | 7/147 (5%) | 8175/30020 (27%) | 73 (49-87) |
| 60-119 days | 8/63 (13%) | 8995/27380 (33%) | 76 (55-88) | 25/165 (15%) | 13727/35572 (39%) | 70 (53-82) |
| ≥120 days | 30/85 (35%) | 11769/30154 (39%) | 60 (38-75) | 85/225 (38%) | 17098/38943 (44%) | 65 (53-74) |

[a] Vaccine effectiveness estimates are not shown against influenza B-associated hospitalizations for some groups due to case counts <50 or confidence interval widths ≥100 percentage points.

[b] Vaccine effectiveness was estimated using logistic regression models comparing the odds of vaccination between influenza-positive cases and influenza-negative controls. Models were adjusted for age, sex, race and ethnicity, calendar day, and site, with age and calendar day treated as natural cubic splines with 4 degrees of freedom.
